# Profiling lockdown adherence and poor coping responses towards the COVID-19 crisis in an international cross-sectional survey

**DOI:** 10.1101/2021.07.21.21260910

**Authors:** Sylvia Van Belle, Amelia Dahlén, Helgi B. Schiöth, Samantha J. Brooks

## Abstract

This study uses international respondents to a COVID-lockdown related questionnaire (n = 1,688) to assess the determinants of adherence and poor coping in response to lockdown measures. A regression analysis was used to compare the relative importance of clusters derived from a K-means cluster analysis as well as various demographics (age, gender, level of education, political affiliation, a factor reflecting social security and a factor reflecting the lockdown harshness). Three distinct clusters (“General Population”, “Extreme Responders” and “Sufferers) were identified, corresponding well to a previous study. Clusters appeared to be the best overall predictors of coping and adherence although gender, political affiliation and lockdown harshness were also important predictors. The large proportion of variance that remains unexplained, combined with the relatively weak effects of traditional demographics, suggest that less concrete variables such as personality traits, health and environmental factors may be better predictors of adherence and coping during a pandemic.

## 1 Introduction

COVID-19, a disease caused by the novel severe acute respiratory coronavirus 2 (SARS-CoV-2), currently represents one of the greatest health challenges posed in the last 100 years [1]. The high infectivity and virulence following its outbreak in Wuhan China in late 2019 led to the declaration of a Public Health Emergency of International Concern on 30 January 2020 by the World Health Organization [2], and the labelling of the disease as a pandemic on 11 March 2020 [3]. At the time of writing in May 2021, nearly 160 million cases of COVID-19 have been identified across 223 countries and territories, and over 3.4 million deaths have been reported [4]. There has therefore been a need to find effective ways to minimise morbidity, mortality and transmission rates.

For this purpose, several COVID-19 vaccines have been developed at unprecedented speeds, and many are viewing vaccines as a silver bullet against COVID-19. Countries such as the United Kingdom (UK) have rapidly descaled all other preventative campaigns [5]. However, synchronised global vaccine roll-out has proved challenging, and the long-term efficacy of vaccination campaigns in modification of pandemic progression through herd immunity is as yet unproven [6].

For this reason, and as a weapon against the emergence of novel threats, behaviour modification remains efficacious as a vital first line of defence [7, 8]. Behaviour modifications have included restricting high-density thoroughfare by closing schools, shops, businesses and imposing travel bans and. Individuals have been advised to frequently sanitise hands and surfaces to reduce fomite-related spread, wear face masks to reduce droplet spread and avoid contact outside their household [8, 9], or, as simply put by the National Health Service [10], to “wash hands, cover face, and make space”.

However, many individuals and businesses have opposed such restrictions [11], with other individual, national, and cultural factors likely impacting adherence to lockdown measures. These factors may also influence psychosocial responses to strict behaviour regulations [12, 13]. Identifying the nature of these determinants may assist in developing campaigns that maximise adherence to regulations while minimising their negative psychological impact.

To that end, several studies have begun asking questions about risk factors that impact individual adherence and well-being during lockdowns, with age and gender commonly highlighted as determinants. For example, older men appeared to implement fewest risk-mitigating behavioural changes early in the pandemic [14, 15], while younger adults have been substantially affected by mental distress and unemployment [16]. Several studies have examined how faith in government relates to pandemic responses. Wong and Jensen [17] reported that increased trust in government related to decreased adherence in Singapore, while Sibley et al. [18] reported that positive attitudes towards government correlated with improved psychological well-being in New Zealand. Geopolitical factors may also play a role, with a survey in the early pandemic of over 116 000 international participants showing that Western European nations had less trust in their governments, and more concern around COVID-19 [19]. Concern and perceived severity of pandemic measures may be another factor related to adherence and coping, as Lieberoth et al. [19] also noted that increased concern was positively related to adherence, but also associated with increased mental distress. However, they also found that increased stress was independently associated with poorer adherence. Perceived severity may interact with factors mentioned above such as age and gender, with Galasso et al. [20] reporting that women showed greater concerns about COVID-19 severity, while younger individuals are known to engage in riskier behaviours [21]. The interaction between perceived risk, adherence and actual disease risk then becomes complicated, as improved adherence may decrease risk of infection. The ambiguity and potential interaction between these risk factors highlight the need for novel methods to clarify the determinants of adherence and coping during a pandemic or other global health crisis.

As an alternative to studying individual risk factors, cluster analysis allows for the grouping of participants according to similarities across multiple dimensions, a technique recently applied to pandemic lockdown response data by Duffy and Allington [22]. They conducted a survey in the UK with 2,250 respondents during the first wave of the pandemic and clustered their participants according to questions relating to adherence behaviours, negative emotions, virus threat perception, trust in leadership and belief in scientifically supported information. The study reported the emergence of three distinct clusters: the “Accepting”, representing 48% of respondents who supported lockdown measures, adhered to guidelines, and were coping well emotionally; the “Suffering”, who made up 44% of respondents, experienced higher levels of anxiety, depression, and compulsive checking of social media; and the “Resisting”, representing 9% of respondents, were unsupportive of lockdown rules and were more likely to disregard government guidance. The study also used demographic questions to further compare the groups, finding that young men were most likely to be “Resisters”[22].

However, Duffy and Allington [22] did not analyse any additional outcomes beyond cluster characterisation. As such, to progress the previous research, this study analyses how clusters and demographics relate to adherence and poor coping during the pandemic. This new cluster analysis is performed on responses to questions similar to those used by Duffy and Allington [22], but in addition reflects an international population that has experienced more months and fluctuating lockdown measures during the pandemic, which may provide new insights. The relationship of demographics such as age, gender, political affiliation and level of education to adherence and coping are examined in the present study, as well as geopolitical factors relating to social security and national pandemic responses. These analyses not only provide data on which groups of people should be targeted by adherence campaigns and provided additional support but may also assist in highlighting which determinants are relevant for further studies of adherence and coping during this and future pandemics.

Given how the ongoing COVID-19 crisis continues to have detrimental effects on medical, economic and mental health dimensions, the current study examines the factors that contribute to lockdown adherence and coping through the following two objectives.

The first aim is to conduct a cluster analysis with an international cohort. The first hypothesis (H1) is that participants will form coherent, statistically significant clusters according to lockdown responses. The second aim is to provide a focused analysis of the impact of the clusters and demographic variables: age, gender, level of education, political affiliation and geopolitical indices on reported adherence to lockdown measures and psychological coping of respondents. The second hypothesis (H2) is that these variables will account for a large portion of the variance in adherence and coping scores.

## 2 Methods

### 2.1 Participant recruitment and ethics

This study draws from an international survey of 2,518 respondents. Self-reported data was collected on Qualtrics, between 27 July and 4 September 2020, in English. Participants were globally recruited through advertisements on social media (i.e. Facebook and Twitter) and Prolific (an on-demand, self-service data collection service) (prolific.co) using a convenience sampling method that provided an international sample. A discussion of the relative advantages and disadvantages, as well as their respondent demographics, are available from Prolific’s website [23]. Participants were incentivised by Prolific’s minimum wage of 1 GBP per hour and the chance to win a £100 Amazon voucher for their participation. Interested respondents were informed about the nature of the study, the steps taken to maintain anonymity and the inclusion and exclusion criteria when they accessed the Qualtrics study webpage.

People were asked to self-exclude if they were under the age of 18, above the age of 75, or had not experienced COVID-19 lockdowns in their country. Participants were asked to provide consent before participating and were given a maximum of one week to complete the survey. Progression locks were applied to the survey to ensure that all tick-box questions were completed, although free-text fields could be submitted empty. Datasets received from Qualtrics were automatically anonymised, although some participants were given the option to provide their email address to enter the prize draw (with the explanation that they would be stored separately for confidentiality). Email data was extracted by the primary investigator into a separate password-protected database which was not shared with other researchers or placed on online servers/cloud storage. Only anonymous data was shared with other members of the research team. The study received ethical approval from the Liverpool John Moores University, UK Research Ethics Committee (UREC ref: 20/NSP/035).

Of the total 2,518 respondents, participants were excluded if they did not complete the survey (n = 828) or reported an age <18 or >75 (n = 2). Participants were excluded for specific analyses if they did not meet the requirements for the test (e.g. missing values). A diagram showing participant selection is shown in S2.

### 2.2 Study design and measures

#### 2.2.1 Demographic information

Demographic information was obtained using three open-ended questions about age, country of residence, highest level of education (School-level exams, undergraduate degree, masters degree, PhD) and two closed questions on gender (man, woman, prefer not to be identified by gender, and other) and political affiliation (far left, left, central, right, far right, or no political interest).

These demographic questions were based on the questionnaire created by [22]. However, highest level of education was included as an additional measure given its association with COVID-19 beliefs [24]. Open-ended questions, particularly “highest level of education”, required extensive codification for standardization which was semi-automated in MySQL using Oracle MyQSL Workbench (v8.0 CE) and custom Python 3 scripts (algorithms available on request). The output was reviewed by two authors (S.V.B and A.D or S.B).

Alongside the demographic variables listed above, geopolitical variables were also examined for a potential influence on coping and adherence. Geopolitical indices were derived from participant country data and response date were used to harvest data from previously published scales, including the “Human Development Index”[25], the Latent Human Rights Protection Score” (also known as the

Human Rights Score (HRS)[26], as well as data on national lockdown severity and disease containment from the Oxford COVID-19 Government Response Tracker (OxCGRT)[27].

#### 2.2.2 COVID Lockdown Questions

Lockdown Questions included fifty-four items assessing behaviours, beliefs, and attitudes around the COVID-19 crisis and national lockdown measures, which were based on publicly available material produced online by Duffy and Allington [22]. The questions used in the survey can be found in S1. Due to the recency of the COVID lockdown, validated tools to measure its impact are lacking, thus it was decided to use tools from prior research to allow for some degree of longitudinal/comparative data. 5-point Likert scales were utilised to indicate agreement with statements covering adherence to official guidance, self-medication efforts (often non-evidence based, e.g. use of antibiotics or ginger tea), beliefs around COVID-19 (e.g. that coronavirus was created in a laboratory), well-being during the crisis/lockdown, non-COVID-19 related health behaviours (alcohol consumption, diet, etc.), beliefs about the future, trust in government (e.g. “I believe the government acted too slowly to control the spread of the virus), and social media usage. Besides the use of a Likert scale, other measures taken to minimise bias included avoiding leading questions, keeping questions short and clear, avoiding difficult concepts and keeping questions simple. Participants who did not complete the survey or had missing answers were excluded, as were statistical outliers were excluded.

Question responses were used to cluster participants based on scores for factors identified by a principal component analysis. The questions were also used to create two dependent variables: 1) an adherence score and 2) a poor coping score. A diagram showing the data processes used can be found in S3. All analyses were run in IBM SPSS (v27.0) and graphs were created in GraphPad Prism (v6.01. Figures were created in Adobe Illustrator.

### 2.3 Variable creation/calculations

The calculation of variables prior to analysis is described below.

#### 2.3.1 Lockdown factors

A principal component analysis (PCA) was run against participant responses to the 54 lockdown-related questions to reduce dimensionality and bring out variances to improve the K-mean clustering. Variance between variables may be diminished due to over-representation of certain dimensions in the survey (e.g. if there were several questions regarding adherence, but only one regarding mood, total variance would be lowered). This is of particular importance in an unvalidated tool [28, 29].

Assumptions of multiple continuous or ordinal variables, linear relationships between variables, sampling adequacy (using a Kaiser-Meyer-Olkin (KMO) measure) and Bartlett’s test of sphericity to determine suitability for data reduction were assessed. The PCA was conducted with responses from 1688 participants using varimax rotation with Kaiser Normalisation and extraction based on Eigenvalues greater than 1. Components (i.e. individual lock-down questions) with a factor loading greater than 0.5 or smaller than -0.5 were considered.

Factors were assessed for coherence/interpretability and “factor variables” were created by inverting the score of components with negative factor loadings (e.g. a score of 1/5 became a score of 5/5), creating a percentage score for each component (e.g. 1 out of 5 became 0.2), and calculating the mean scores for the components within the variable, to prevent loading in the cluster analysis. Factors were then multiplied by the median “highest possible score” for the components (typically a score of 5, except for the final factor, which consisted of a single Boolean question, and was thus scored out of 2) to provide more interpretable scores.

#### 2.3.2 Clusters

To evaluate H1 and provide clusters as a variable for H2, a K-Means cluster analysis was performed in line with Duffy and Allington [22]. The current analytical method differed from the reference study by implementing dimensional reduction through a principal component factor analysis of responses to the lockdown questions to improve the robustness of the K means clustering as dimensional reduction through a factor analysis of the lockdown question responses improves the robustness of the K-means-clustering.

Following the dimensional reduction, serial K-Means cluster analyses using varimax rotations with Kaiser normalization were run on the same participants using the created factor variables. To determine the optimal number of clusters, cluster analyses were run for 2 <= k <= 10, where k represents the number of clusters, saving cluster membership for each participant for each analysis. A cluster would be considered valid if the analysis stably converged within 30 iterations. The sum of squared errors for each analysis was plotted to visually determine an optimal k value at an “elbow” point, and an exploratory ANOVA was run for each of the clusters in each analysis against the factor variables for evaluation of coherence/interpretability.

Once an optimal k had been determined, a formal analysis of variance was run against the lockdown factors used for the cluster analysis, to clearly describe the variance between them. The distribution of the data, evaluated with a Kolmogorov-Smirnov test, determined whether a MANOVA and t-tests (for normally distributed data) or Multivariate Kruskall-Wallis (MKW) tests with eta-squared values were performed (for non-parametric distributions). Outliers with a Mahalanobis value >= 31.264 (the critical value for the 12 lockdown factors included in the analysis) were excluded. Log-transformation would only be used if it resulted in improved distribution. A variance inflation factor (VIF) was used to assess collinearity between dependent variables (i.e. lockdown factors) and a Levene’s test was used to assess homoscedasticity.

The demographics of the clusters were also assessed: Chi squared tests were used to evaluate how gender, politics and education relate to the participant clusters, while a Kruskal Wallis one-way ANOVA was run for continuous demographics (age, and geopolitical factors), checking assumptions of independence of observations, a linear relationship, normal distribution, absence of multicollinearity and homoscedasticity, using eta-squared values as a measure of effect size.

#### 2.3.3 Demographic factors

To reduce the number of dimensions included in the final analyses, assumptions were assessed and an additional PCA was run according to the same method described above (Section 2.3.1), now using the participant scores for the various geopolitical indices: country-matched Human Rights Score (HRS) and Human Development Index (HDI) score, and four indices from the OxCGRT measures (government response, containment and health, economic support and lockdown stringency) matched to country and participant questionnaire submission.

Summarised geopolitical scores were created as a fraction of 1 (OxCGRT scores were represented as a percentage and were thus divided by 100, HDI was already a score out of 1, and HRS ranged between ±4, and so 4 was added and then scores were divided by 8). Scores were omitted for participants with missing data, either due to participants not providing a valid country (n = 4), or due to missing data in the indices for the provided country or date (n = 122).

#### 2.3.4 Outcome measures: adherence and coping

A score for adherence, with a maximum of 60, was calculated by summing scores for the following 12 questions:

I followed the COVID lockdown rules

I supported lockdown measures

I am adhering to social distancing rules

I wash hands more often for at least 20 seconds

I cover my mouth when coughing

I avoid close contact with someone who is infected

I have been avoiding places where many people are most likely to gather (e.g. parks, beaches, other outdoor spaces)

During the COVID lockdown I met up with friends or family outside the home (inversely scored)

During the COVID lockdown I have had friends or family visit me at home (inversely scored)

I have been outside when having coronavirus-like symptoms (inversely scored)

I agree with the following statement: “The NHS recommends that you should wear a face mask when you are out, even if you do not have coronavirus”:

I am closely following official guidance/recommendations on how to protect myself and others

A score for poor coping with a maximum score of 61 was calculated by summing scores for the following 13 questions:

I have been in contact with a counselling or support service

I have lost sleep over coronavirus

My eating patterns have changed during the coronavirus lockdown (0 = eating patterns have not changed, 1= eating more or eating less than usual)

I am eating much more UNHEALTHY food

I am drinking much more alcohol

I am using non-prescription drugs (e.g. painkillers, other over-the-counter remedies) much more

I am finding the coronavirus outbreak and/or the lockdown measures extremely difficult to cope with

I have been spending time thinking about the coronavirus

I have argued more with family/people in the home during the COVID lockdown

I feel more anxious since the lockdown measures were introduced

I feel more depressed since the lockdown measures were introduced

I feel helpless as a result of coronavirus

I check social media for information or updates about coronavirus:

(total possible score of 61)

### 2.4 Determinants of adherence and poor coping

A hierarchical multiple regression was performed for both outcome variables to create a model of their determinants and to understand how demographic variables and clusters account for variance in respondents’ adherence and poor coping scores. A hierarchical regression allowed the combined effect of education, political affiliation and calculated demographic factors to be determined, after the combined effect of age and gender has been accounted for. Similarly, the amount of variance explained by clusters once the effects of the other demographic variables have been accounted for could then also be evaluated. This hierarchical approach allows for interactions between independent variables. A diagram showing the regression hierarchy is presented in S4.

The significance, but not effect size, of individual variables used in each model was provided by the regression analysis. For an approximations of effect size, individual multivariate Kruskal-Wallis tests or linear regressions were used for each variable against the outcome variables. Percent change between medians of categorical variable scores was calculated and percent change between the extremes of linear variables as interpolated from the regression line formula was calculated for easy comparison between different measures. It should be noted that these individual tests do not account for the effects of covariates and should thus be interpreted alongside the P-values for variables provided by the regression analysis.

## 3 Results

### 3.1 Variable creation

#### 3.1.1 Principal component analysis for lockdown questions

A PCA was run on responses to 54 lockdown-related questions, using n = 1,688 as described in S2 and inspection of the correlation matrix (available on request) showed that all variables had at least one correlation coefficient greater than 0.3. The overall Kaiser-Meyer-Olkin (KMO) measure was 0.886, classifying as ‘meritorious’ according to Kaiser [30]. Bartlett’s test of sphericity was statistically significant (p < .001), indicating that the data was likely factorizable.

The PCA revealed 12 components, which cumulatively explained 56.43% of the total variance. The full rotated component matrix is available in S5. Questions were deemed relevant for the factor if they had factor loading > 0.5 or < −0.5. The factors were coherent with similarly themed questions, and were termed as follows: self-treatment, government response, poor mood, adherence, fringe beliefs, optimism, unhealthy consumption, in-person contact, lockdown expectations, indirect transmission, financial insecurity and suspected infection. A list of the questions included in each factor can be found in S6.

There were no questions with sufficient loading for inclusion in multiple factors. Questions that had insufficient factor loading (−0.5 < x < 0.5) were omitted from the factor variables, and question 15 (“I have been outside when having coronavirus-like symptoms) was excluded from the factor “self-medication” due to lack of interpretability/coherence with the other questions included in the factor.

#### 3.1.2 K-means clustering

K-means cluster analyses were performed to examine whether clear subgroups were present in the population with respect to their responses to the factors described in the PCA above. The optimal number of clusters had to be determined, that iterative K means analyses were run between using two to ten clusters. The sum of square residuals for each analysis with cluster number = *k* did not resolve into a clear “elbow” to suggest an optimal number of clusters when plotted, however, clusters only resolved within 30 iterations for *k* = 2, 3 and 6, and it was therefore decided to analyse only *k* = 3 for comprehensibility and compatibility with Duffy and Allington [22], and to maintain a meaningful population size within each cluster.

##### Cluster composition

To describe and differentiate these three clusters, the distributions of participant scores within each cluster were compared for each of the thirteen lockdown factors. The distributions of scores for the lockdown factors within the clusters were all significantly skew. Distribution was not improved by excluding outliers (n = 27, Mahalanobis value >= 31.26), or through log transformation (data available on request). Data was used without log transformation to maintain interpretability, but outliers were excluded from cluster comparisons and further cluster-based analyses (n = 1,661).

A non-parametric MKW was run with the complete results available on request. Briefly summarised, 51 out of 66 factor comparisons between clusters had significantly different scores. Interactions that were ***not*** significant (i.e. *p* > 0.05) included *Government Response* between Cluster 1 and Cluster 2 (*p* > 0.999), *Adherence* between Cluster 1 and Cluster 3 (*p* = 0.250), *Fringe Beliefs* between Cluster 1 and Cluster 3 (*p* = 0.110), *Poor Mood* between Cluster 2 and Cluster 3 (*p* = 0.282), and *Lockdown Expectations* between Cluster 2 and Cluster 3 (*p* = 0.396).

Cluster 1 (n = 820) had the lowest score for *Poor Mood*, the least *Unhealthy Consumption*, and was the least *Financially Insecure*. This cluster was therefore labelled the “General Population” as there were no other discriminators.

Cluster 2 (n = 211) scored highest on *Self-Treatment* and *Fringe Beliefs*, while also reporting more *In-Person Contact, Poor Mood* (compared to Cluster 1), and *Indirect Transmission*. Conversely, this cluster had the most *Optimism* and worst *Adherence*. This cluster was therefore labelled “Extreme Responders”.

Cluster 3 (n = 630) had *Poor Mood* compared to Cluster 1, and also had the worst *Financial Insecurity* as a result of the pandemic. They had the lowest *Optimism* and *Government Response* scores. This cluster was therefore labelled “Sufferers”.

#### 3.1.3 Demographics principal component analysis

A PCA was run on participant scores (n for the six continuous geopolitical indices: the HDI (based on participant country), HRS (based on participant country), and the four OxCGRT variables (based on participant country and the date of questionnaire submission). To assess assumptions, inspection of the correlation matrix (available on request) showed that all variables had at least one correlation coefficient greater than 0.3. The overall Kaiser-Meyer-Olkin (KMO) measure was 0.583, classifying as ‘miserable’, but not unacceptable, according to Kaiser [30]. Bartlett’s test of sphericity was statistically significant (p < .001), indicating that the data was likely factorisable.

The PCA revealed two factors that had eigenvalues greater than 1, which cumulatively explained 88.98% of the total variance. Variables were highlighted as relevant for the factor if they had a loading of > 0.5 or < -0.5. The rotated component matrix can be found in S7. The factors were coherent with similarly themed variables. The factor subsequently termed the “National Coronavirus Response Factor” (NCRF) would incorporate OxCGRT scores for government response; containment and health polices; and lockdown stringency. Participant scores ranged from 0.17 to 0.48. The factor subsequently termed the “Social Security Factor” (SSF) would incorporate the Human Rights Scale, a measure of protection of physical integrity; the Human Development Index, a measure of life expectancy, years of schooling and gross national income; and the OxCGRT score for economic support during the pandemic. While economic support during the pandemic loaded above 0.5 for both factors, it was decided to include it in the calculation of SSF, as it loaded highest for this factor in the PCA. Participant scores for the SFF ranged between 0.07 and 0.80.

### 3.2 Descriptive statistics

#### 3.2.1 Population Demographics

An overview of respondent population (n = 1,688) demographics can be found in Fig. 1.

**Fig. 1:**
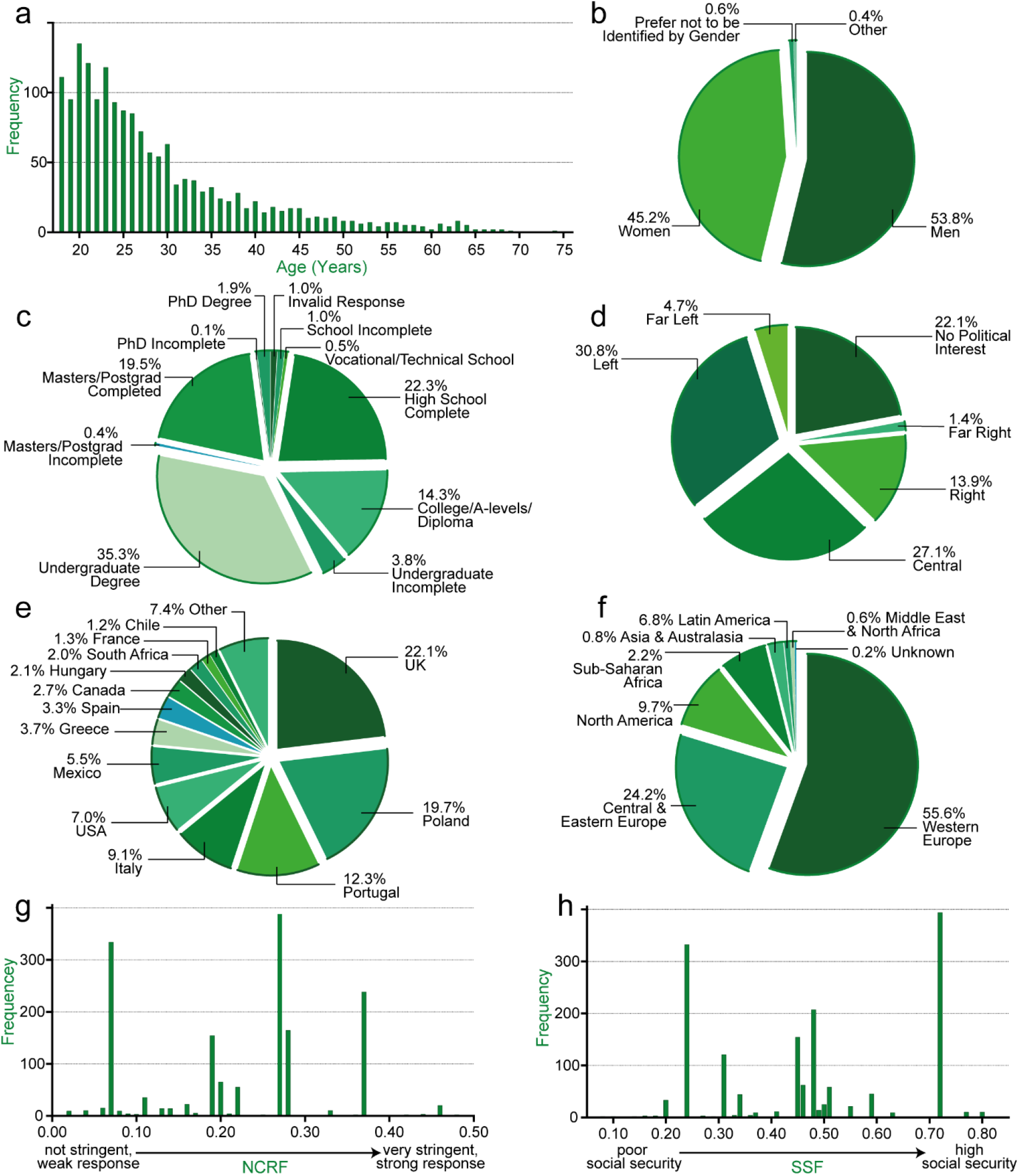
Participant distributions for demographic variables. (a) frequency histogram showing participant age distribution. (b) pie chart of respondent gender proportions. (c) pie chart of proportional level of education using author interpretations of free-field text responses. Postgrad = postgraduate. (d) pie chart of proportional political affiliation reported by respondents. (e) pie chart of participant nationalities (f) pie chart of participants by international regions (g) frequency histogram showing distribution of participants’ calculated National Coronavirus Response Factor (NCRF) scores, a measure of COVID-related lockdown stringency, government response and virus containment. (h) frequency histogram showing distribution of participants’ calculated Social Security Factor (SSF) scores, a measure of country development and human rights protection, as well as economic support provided during the pandemic.

Ages of respondents were significantly skewed, as 50.65% of respondents were 25 or younger (n = 855).

There was a slight skew towards men (53.8%, n = 908) compared to women (545.2%, n = 762), and an insufficient sample of non-binary respondents who selected either “prefer not to be identified by gender” or “other” (1.06%, n = 18) to include in comparative analyses, and thus unfortunately had to be excluded.

Political affiliation was treated as ordinal data with “central” scoring halfway between “far left” and “far right” and respondents with “no political interest” were excluded from further analyses. Responses were distributed amongst left (30.8%, n = 519), central (27.1%, n = 457), no political interest (22.1%, n = 373) and right (13.9%, n = 234). Extremes only accounted for 6.2% of the responses (n = 105).

Level of education was also treated as ordinal data, categorised from a free-text field according to authors’ discretion (algorithm available on request). 22.3% of respondents (n = 376) reported that they had only completed high school, 14.3% (n = 241) had studied as far as college/A-levels/diploma level studies, 35.51% (n = 596) had completed an undergraduate degree and 19.5% had completed a masters or equivalent postgraduate degree (n = 328). Those who gave invalid/uninterpretable responses (n = 16) were excluded from further analyses.

#### 3.2.2 Cluster Demographics

Differences between demographic variables for each cluster, along with cluster composition for each lockdown factor, are shown in Fig. 2. Participant with invalid values for demographic variables were excluded, resulting in n = 1,221, as described in S2.

**Fig. 2:**
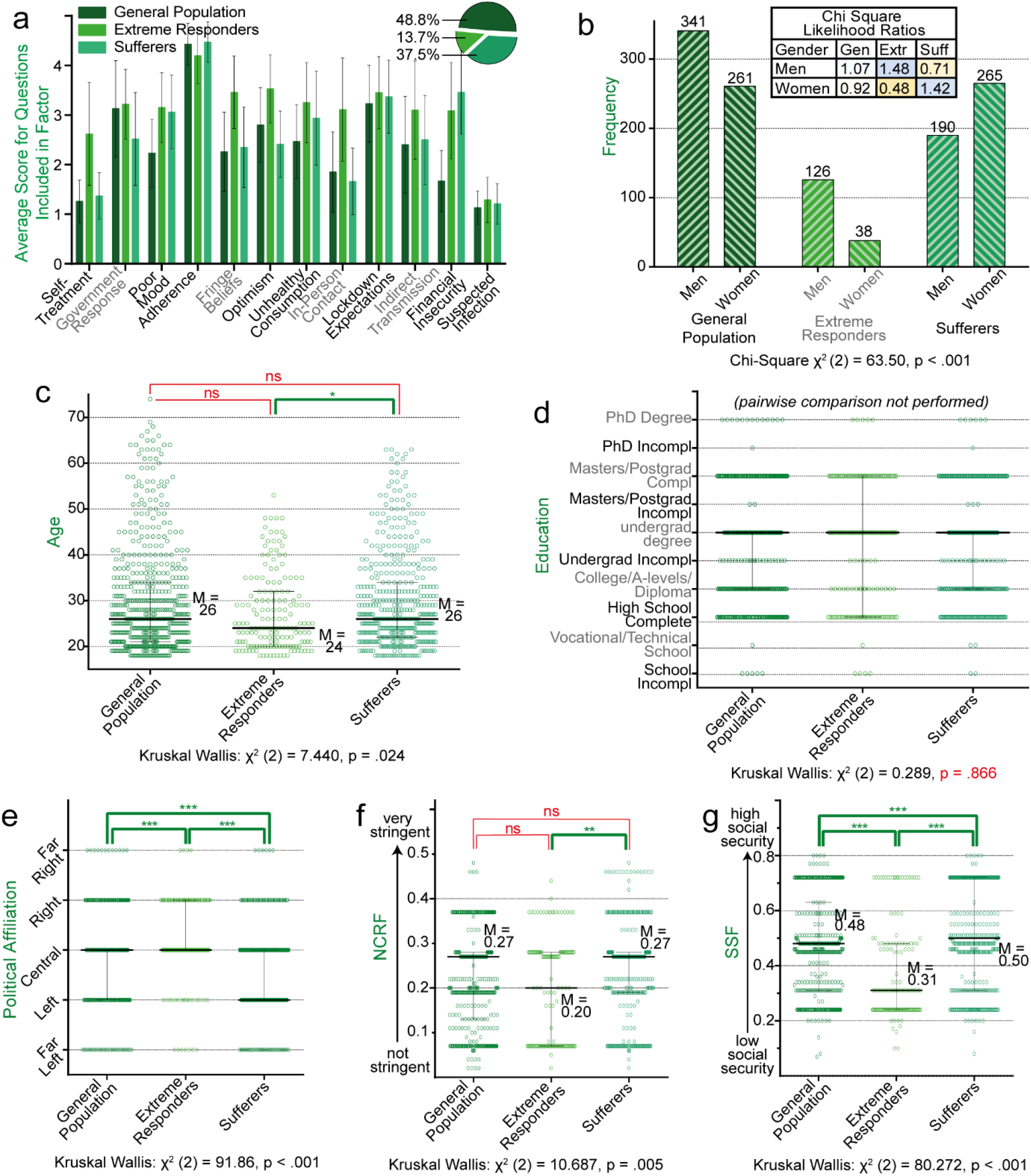
Participant cluster composition. (a) Bar graph showing cluster scores for each lockdown factor, with proportion of respondents in each cluster shown in the top right. Participant scores for each factor are their average score for each question included in the factor out of 5. The factor “suspected infection” is a binary score where 2 = suspected infection and 1 = infection not suspected. (b) Frequency histogram showing the number of men and women in each cluster. Results from a Chi-squared test are stated below the graph and the likelihood table reflects the likelihood that a participant of a particular gender will be in a specific cluster. (d-g) Grouped scatter plots showing the distribution of respondents’ demographics for each cluster. Results of multivariate Kruskal-Wallis tests are stated below each graph, and Dunn-Bonferroni post-hock comparisons were used to test for between-cluster significance. Except for (a), which shows mean and standard deviation, error bars show the median and interquartile range. Gen = General Population cluster; extr = Extreme Responders cluster; suff = Sufferers cluster; n.s. = not significant; M = Median; -grad = -graduate; incompl = incomplete; SSF = social security factor; NCRF = national coronavirus response factor. *** = p < 0.001; ** = 0.001 ≤ p < 0.01; * = 0.01 ≤ p ≤ 0.05; n.s. = p > 0.05.

A chi-squared test showed that there was significantly different gender distribution between clusters. Women were approximately half as likely as men to be extreme responders (Likelihood ratio = 0.482) less likely to be extreme responders, and more likely to be sufferers (Likelihood ratio = 1.424).

MKW tests for ordinal and linear variables showed significant between-cluster differences for age, political affiliation, the SSF and the NCRF, but not for level of education. Post-hoc Dunn-Bonferroni between-group comparisons, showed that Sufferers were slightly older than Extreme Responders, tended to come from countries with a higher NCRF than Extreme Responders and were the most left-leaning of the clusters. Extreme Responders had the lowest SSF and were right-leaning politically with a lower NCRF than Sufferers. Further details of the pairwise comparisons are available upon request.

### 3.3 Determinants of adherence and poor coping

#### 3.3.1 Adherence

Results from a hierarchical multiple regression of demographics and clusters against adherence scores to determine relative contribution of each determinant to the outcome are shown in Fig. 3a. Assumptions are reported on in the S8.

**Fig. 3:**
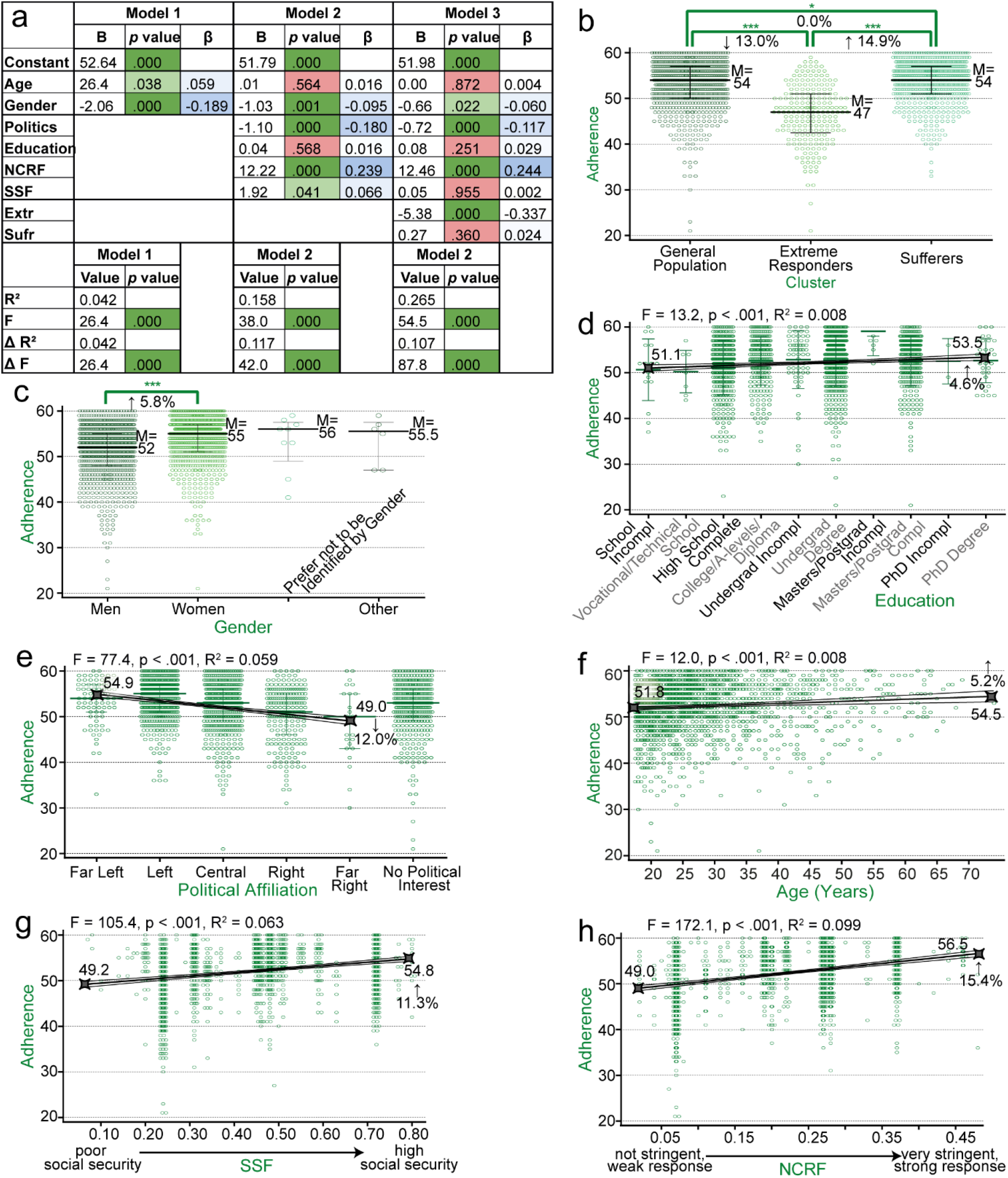
Determinants of adherence. (a) Summary of a hierarchical regression for the relative importance of the determinants of adherence. β = standardised correlation coefficient for comparison of between variables. The R2 is equivalent to the variance explained by the model. (b-h) Individual analyses to evaluate how each variable explains variance in adherence. (b,c) Grouped scatter plots for clusters and gender showing medians (M) and interquartile ranges for adherence scores. Significance for each comparison, determined through Kruskal Wallis tests, is indicated by bracketed asterisks and the relative change (from left group to right group) is indicated as a percentage. (d-h) Scatter plots for adherence scores for level of education, political affiliation, age, the national coronavirus factor (NCRF) and social security factor (SSF) with regression lines and 95% confidence bands plotted. The statistics for the regressions are stated above each graph. Minimum and maximum values for the regression lines were calculated and the direction and relative change are shown as an indication of effect size.

The full model using demographic factors and clusters as predictors for adherence was statistically significant (*p* < .001). Age and gender (model 1) explained 4.2% of the variance in adherence (i.e. had an R^2^ of .042), while the addition of politics, level of education, NCRF and SSF explained a further 11.7% of the variance. The addition of clusters explained a further 10.7% of the variance.

The standardised coefficients (β), which show variation of adherence in terms of standard deviations of the independent variables for comparison between elements, highlight that being within the extreme responder cluster, the NCRF, political affiliation and gender are, in order, the important determinants of adherence, while age, level of education, the social security factor score, and the Suffering cluster did not correlate significantly with adherence scores in the final model (*p* values of .872, .251, .955 and .360 respectively).

When cluster was not included in the model (model 2), 15.8% of the variance was explained, mainly through the NCRF, political affiliation, and gender.

Due to the skewed distributions within subgroups, formal determination of effect size including the effects of covariates would be subject to high chances of error. To provide an idea of the direction and magnitude of the interactions between individual variables and adherence, individual regressions or multivariate Kruskal Wallis tests with Dunn-Bonferroni post-hoc tests were run (Fig. 3b-h), and although these tests do not account for the effects of covariates, they provide insight into how each variable.

Agreeing with the regression model, individual tests showed that Extreme Responders were significantly likely to have worse adherence scores than both the General Population (eta^2^H = 0.282, *p* < .001) and Sufferers (eta^2^H = 0.324, *p* < .001) and men were more likely to have worse adherence scores than women (eta^2^H = 0.022, *p* < .001). Adherence tended to decrease as political affiliation tended towards right-ness (*p* < 0.001), and was positively correlated with NCRF (*p* < 0.001) – i.e. countries with harsher lockdowns and better containment during the crisis had better adherence scores.

Although non-significant in the overall regression model, individual analyses showed a statistically significant interaction between adherence and age (*p* < 0.001), level of education (*p* < 0.001) and SSF (*p* < 0.001).

Effect sizes were typically small across tests. For more interpretable measures of effect size, the regression line formula was used to interpolate minimum and maximum scores for each independent variable. Across ages 18 to 75, there was an increase in adherence scores of 5.2%; from those who affiliated with the far-left politically to those who associate with the far-right, there was a decrease in adherence scores of 12.0%; and from those who had not finished high school to those who had a PhD degree there was an increase of 4.6%. From the lowest to the highest calculated NCRF scores, there was an increase in adherence scores of 15.3%; and from the lowest to the highest calculated SSF scores, there was an increase of 11.3%. The median score for adherence was 5.8% higher for women than for men, while for clusters, the median score for extreme Responders was 13.0% lower than both the General Population and Sufferers, who had the same adherence scores.

#### 3.3.2 Poor coping

Results from a hierarchical multiple regression of demographics and clusters against poor coping scores are shown in Fig. 4a. Assumptions are again reported in S8.

**Fig. 4:**
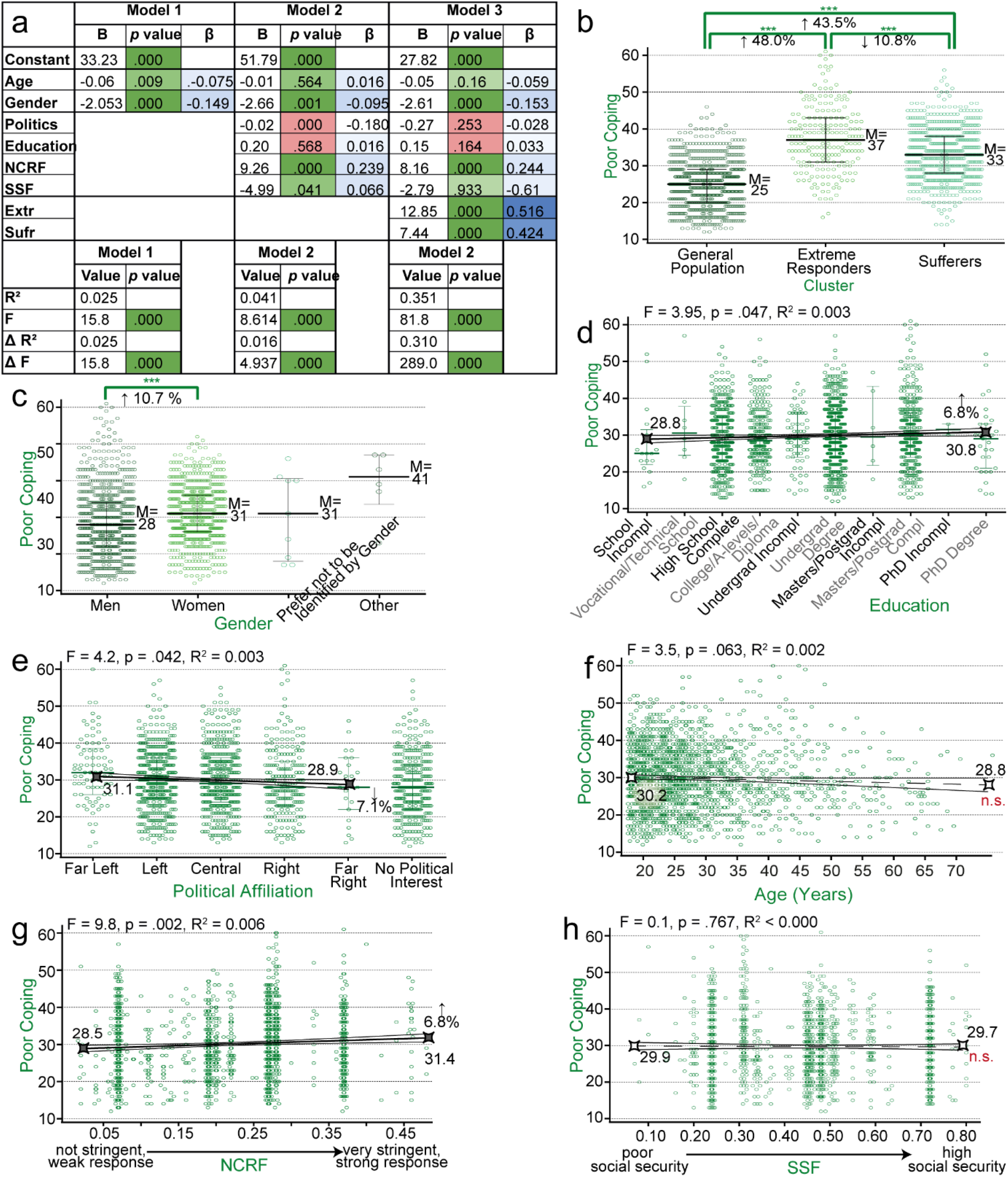
Determinants of poor coping. (a) Summary of a hierarchical regression for the relative importance of the determinants of poor coping. β = standardised correlation coefficient for comparison of between variables. The R^2^ is equivalent to the variance explained by the model. (b-h) Individual analyses to evaluate how each variable explains variance in poor coping. (b,c) Grouped scatter plots for clusters and gender showing medians (M) and interquartile ranges for poor coping scores. Significance for each comparison, determined through Kruskal Wallis tests, is indicated by bracketed asterisks and the relative change (from left group to right group) is indicated as a percentage. (d-h) Scatter plots for poor coping scores for level of education, political affiliation, age, the national coronavirus factor (NCRF) and social security factor (SSF) with regression lines and 95% confidence bands plotted. The statistics for the regressions are stated above each graph. Except in regressions that were not significant, minimum and maximum values for the regression lines were calculated and the direction and relative change are shown as an indication of effect size. n.s. = not significant.

The full model using demographic factors and clusters as predictors for adherence was statistically significant (*p* < .001). Age and gender (model 1) explained 2.5% of the variance in poor coping (i.e. had an R^2^ of .025), while the addition of politics, level of education, NCRF and SSF explained a further 1.6% of the variance. The addition of clusters explained a further 31.0% of the variance.

The standardised coefficients (β) highlight that cluster, gender and NCRF are, in order, the important determinants of poor coping. Politics and level of education did not correlate significantly with poor coping in the final model (*p* values of .253 and .164, respectively).

When cluster was not included in the model (model 2), only 4.1% of the variance was explained, mainly through gender, national coronavirus response factor, the SSF, and age, in order of effect on poor coping scores.

As above, to determine the individual interactions between each determinant variable and poor coping, distributions were graphed and individual regressions and multivariate Kruskal Wallis tests with Dunn-Bonferroni post-hoc tests were run to explore how each of the demographic factors included in the model above relate to poor coping scores (Fig. 4b-h).

Agreeing with the regression model, individual tests showed that Extreme Responders were significantly likely to have poorer (i.e. higher) coping scores than both the General Population (eta^2^H = 0.651, *p* < .001) and sufferers (eta^2^H = 0.324, *p* < .001), and sufferers had poorer coping scores than the General Population (eta^2^H = 0.331, *p* < .001). Women were more likely to have a higher score for poor coping than men (eta^2^H = 0.036, *p* < .001). Poorer coping was positively correlated with NCRF (*p* < 0.001) – i.e. participants from countries with harsher lockdowns and better containment during the crisis had poorer coping scores.

Agreeing with the combined model, the individual regression of SSF against poor coping scores was not significant (*p* = .767). Unlike the combined model, however, the individual regression for age did not deem the relationship with poorer coping significant (*p* = .063), and political affiliation was seen as significant (*p* = .045)

Effect sizes again were typically small. From the lowest to the highest calculated NCRF scores, there was a worsening in coping scores of 10.0% and the median coping score of women was 10.7% worse than that of men. Extreme Responders had a median poor coping score that was 48.0% worse than that of the General Population and 10.8% worse than Sufferers. The median coping score of Sufferers was 43.5% worse than that of the General Population.

Effect sizes were not calculated for age and SSF as the regression was determined not to be significant, reflected by small β coefficients in the multiple regression. Political affiliation and level of education, deemed not to have a significant effect on the overall model, showed a significant effects when evaluated in isolation: from those who affiliated with the far-left politically to those who associate with the far-right, there was an improvement in their coping scores of 7.1%; and from those who had not finished high school to those who had a PhD degree there was a worsening of coping scores by 6.8%.

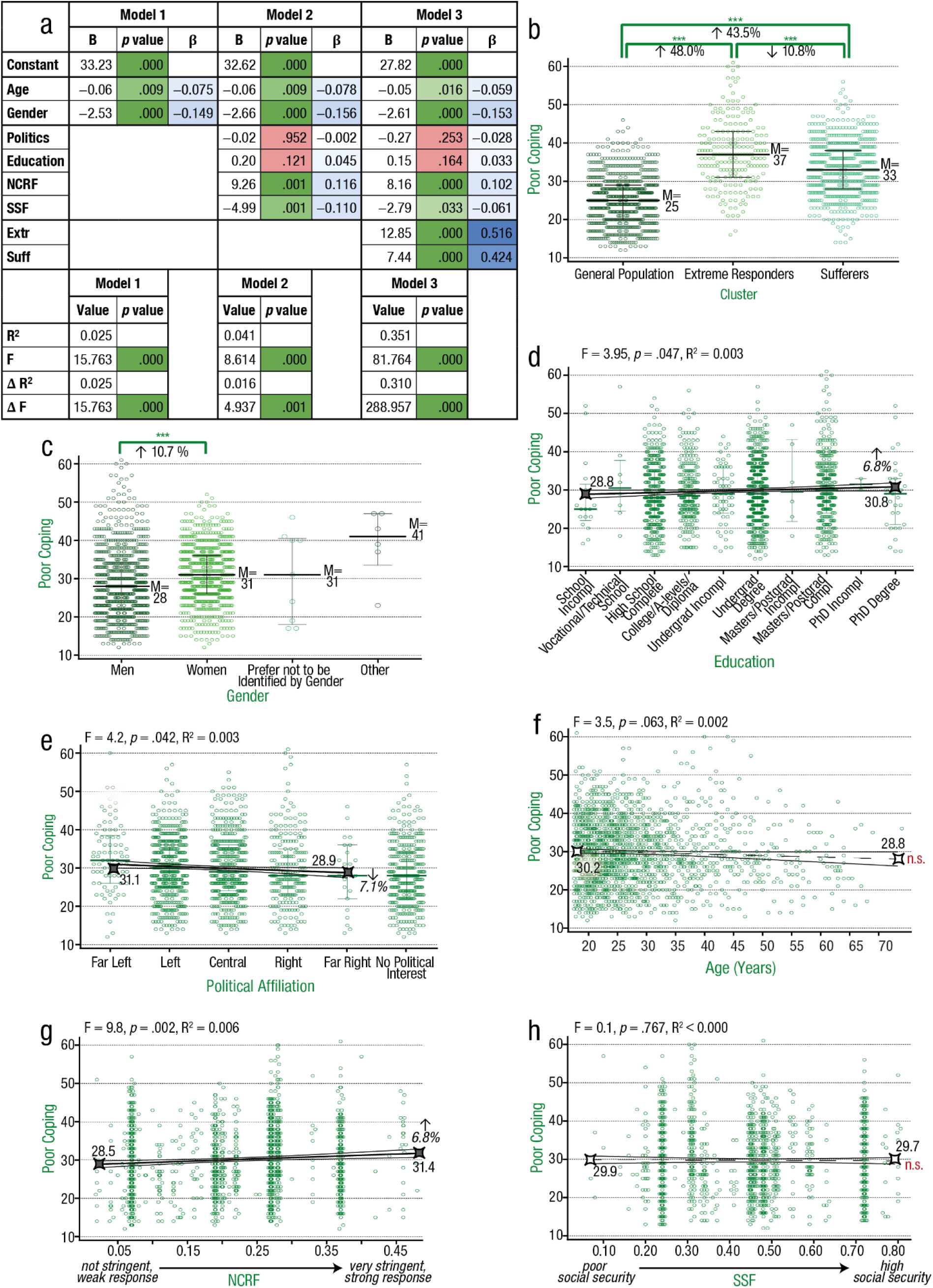

## 4 Discussion

This study evaluated determinants of adherence and poor coping in response to lockdown regulations during the COVID-19 pandemic between July and September 2020. The three clusters, derived from a cluster analysis, accounted for a large portion of the variance in both adherence and poor coping scores (Fig. 5). Other demographic variables also explained a large amount of the variance in scores, particularly gender and lockdown stringency. When analysed individually, the Sufferers cluster (37.5% of responders) and Extreme Responders cluster (13.7% of responders) reported significantly worse coping scores compared to the General Population cluster (48.8% of responders), while Extreme Responders were significantly worse at adhering to lockdown measures in comparison to the other two clusters (Sufferers and the General Population had the same median adherence scores). Women showed higher adherence scores and worse coping scores than men, and participants from countries with harsher lockdowns reporting increased adherence. These major findings and their practical implications will be discussed in turn.

**Fig. 5:**
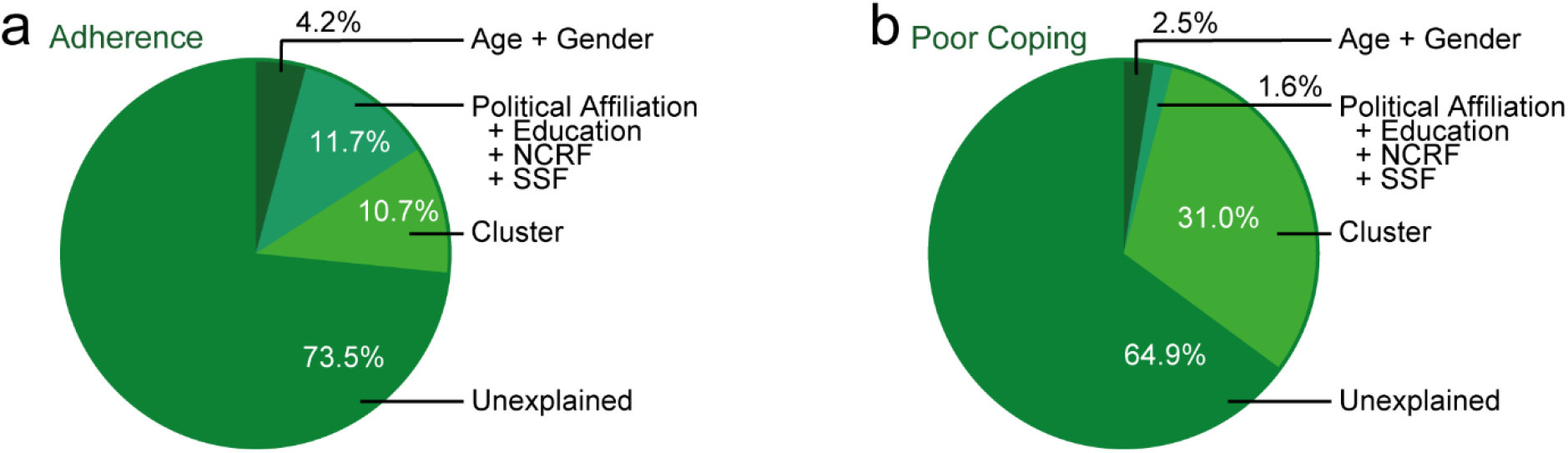
Pie charts showing how determinants contribute to variance in adherence (a) and poor coping scores (b). NCRF = national coronavirus response factor. SSF = social security factor.

The combined predictive value of gender and age on adherence, and, to a lesser extent, poor coping, was significant. However, most of the combined gender and age variance in adherence and coping could be explained by differences across gender rather than age. Women had poorer coping scores, while men had weaker adherence to government lockdown restrictions, which is in accordance with the global trends observed during the pandemic [20, 31]. Alarming reports from China, South Korea,

Japan, Italy, UK and USA estimated that women were 24% more likely than men to permanently lose their job during the pandemic [32]. Therefore, women’s psychological distress during COVID-19 may be linked to greater career and financial insecurities, which could underlie their increased likelihood of belonging to the Sufferers [32, 33]. The reduced adherence seen among men could be due to lower perceptions of social responsibility and higher self-interest [34].

In contrast to gender, the effect of age alone was very small for poor coping and was not significant for adherence. The lack of effect of age may relate to the skewness of the sample; 74.5% of respondents were between 18 and 32 years of age. However, if the lack of effect is a true finding, this may aid in understanding the inconsistency of results from previous studies. Tull et al. [35] found that age had no influence on stress, however Pearman et al. [36] found increased anxiety in older populations and yet Nwachukwu et al. [37] found increased stress, anxiety and depression in young adults. We speculate that there are differing risk factors for poor coping across various ages. Younger adults may have less structural, financial and emotional resilience [16, 38] and struggle with interrupted studies or finding early-career opportunities. Older adults, on the other hand, may have increased anxiety due to their higher risk for severe/fatal COVID-19 and increased isolation resulting from less familiarity with online communication platforms. While there is less information on the relationship between age and adherence, media and government spokespeople have alternately attributed poor adherence to the party-attending, invincible young and the inflexible older adults who think that mask wearing is an infringement on their personal freedom. It should be borne in mind that there may not be clear risk groups across age, but that within each age group, there may be specific risk factors for poor adherence or coping that should be identified and addressed by researchers, policy makers and communicators.

Even after the variance explained by age and gender had been accounted for, the additional variance explained by the combined remaining demographic variables still contributed significantly to variance in adherence and, to a very small extent, poor coping scores. These demographic variables include education, political affiliation, the NCRF (based on country- and date-specific OxCGRT data on lockdown severity and disease containment for each country), the SSF (based on the HRS, HDI and OxCGRT measure of pandemic-related economic support).

Of these four variables, the NCRF provides the largest portion of the variance. To our knowledge, this is the first study to directly compare lockdown stringency with adherence and poor mental health, showing elevated adherence scores but poorer coping in countries with stricter COVID-related regulations, although Lieberoth et al. [19] related OxCGRT Stringency Index scores to individuals’ trust in their governments. Interestingly, in our study, when adherence and poor coping were directly compared, no significant correlation was established (*p* = .063), suggesting that lockdown stringency is an independent mediator of these outcomes. This highlights the importance of finding a balance between the stringency of lockdown and the mental health of a populace.

By contrast, there was no significant relationship between political affiliation and poor coping, although there was a correlation in the final model showing that adherence scores decreased as participants became more right-leaning. This may reflect a belief that lockdown regulations are an infringement on individual freedoms [39, 40].

Lower levels of education have similarly been proposed to contribute to weaker adherence, potentially due to inabilities to work from home and other adversities related to lower socioeconomic status [41, 42]. However, in this study, level of education was unable to reliably predict either adherence or poor coping. It is important to note that the free-text responses for education level limited accuracy as they required subjective interpretation for codification and that the role of education as a predictor for adherence and coping may vary across countries.

The SSF, the last of the demographic variables included in this model, was again not a significant determinant for adherence and had a near-zero effect on poor coping – an interesting finding, considering that environmental social security is often linked with mental health outcomes [43, 44].

Our derived social security factor based on nationality may not be sufficiently sensitive to capture any variability caused by uncertain living circumstances and access to basic human rights.

Finally, as the third and last model added the regression analysis, clusters still explained 10.7% of variance in adherence scores and 31.0% of variance in poor coping scores, even after the effects of demographics had been accounted for. In individual analyses, which do not incorporate interactions across covariates, Extreme Responders had significantly worse adherence and coping scores than the other two clusters. Sufferers had worse coping scores than the General Population, but slight better scores than Extreme Responders, and the same adherence scores as the General Population. Considering the cluster attributes, it could be suggested that Sufferers attempt to offset poor coping by remaining adherent, despite a lack of faith in government responses to the pandemic and a more negative view of the near future. Conversely, Extreme Responders attempt to offset poor coping by exploring alternative theories and explanatory models, disregarding government regulations and minimising the seriousness of the pandemic.

The above cluster-related findings are strengthened by similarities in cluster composition to clusters found by Duffy and Allington [22] in their study. They identified three distinct clusters which they termed the Accepting, Resisting and the Suffering. That said, questionnaire differences and differences in statistical analyses limit direct comparison, specifically their use of UK respondents early in the pandemic compared to our use of international participants several months later, their use of discrete responses (typically binary, reporting results as “% of population who engage in behaviour x”) compared to the Likert-scale responses utilised in this survey. Their study reported findings for each individual question, whereas our study utilised a PCA analysis to provide more focused report – therefore questions best approximating our clusters were used as a surrogate for comparison between the two studies, as shown in Table 1.

**Table 1:** A comparison of cluster findings from Duffy and Allington (2020) and cluster findings from the current study. All comparative words used in the table refer to comparisons with the Accepters/General Population values.

|  | Duffy & Allington (2020) | This study |
| --- | --- | --- |
| Accepters/ General Population | 48% of responders | 49% of responders |
|  | 59% men | 57% men |
|  | Mean age 50 | Median age 26 |
| Sufferers (versus Accepters/General Population) | 44% of responders | 37% of responders |
|  | 64% women | 58% women |
|  | Mean age 44 | Median age 26 |
|  | 27.5% fewer people voted Conservative and 42.9% more people voted Labour | 9.7% more left leaning across cluster participants |
|  | 11.0% fewer people trusted government to control the spread of COVID-19 | 19.5% lower scores for faith in their governments' responses |
|  | No difference in the number of people believing in a quick return to normality | 13.9% lower scores for positive lockdown expectations |
|  | 67.9% more people reported that they were likely to face financial difficulties | 107.2% higher scores for likelihood to face financial insecurity |
|  | 6.9% more people followed lockdown measures | No difference between adherence scores. |
|  | 1062.5% more people reporting anxiety/depression | Had 32.0% worse coping scores |
| Resisters/Extreme Responders (versus Accepters/General Population) | 9% of responders | 13% of responders |
|  | 64% men | 77% men |
|  | Mean age 29 | Median age 24 |
|  | 45.0% fewer people voted Conservative and 66.7% more people voted Labour | 16.9% more right leaning across cluster participants |
|  | 1.4% fewer people trusted government to control the spread of COVID-19 | 2.9% higher scores for faith in their governments' responses |
|  | 200% more people believed in a quick return to normality | 26.1% higher scores for positive lockdown expectations |
|  | 132.1% more people reported that they were likely to face financial difficulties | 85% higher scores for likelihood to face financial insecurity |
|  | 43.7% fewer people followed lockdown measures | 13% lower adherence scores |
|  | 550.0% more people reporting anxiety/depression | 48.0% worse coping scores |

From this table, the most notable similarity between the two studies is the participant and gender distribution across clusters. We were unable to replicate the age differences between clusters due to a very skewed age distribution of our respondents (median age 26). Sufferers from both studies were more left-leaning, although Duffy and Allington [22] found that their “Resisters” were more left-leaning than their “Accepters”, in contrast to our Extreme Responders, who were more right-leaning than our General Population. Both studies reported increased financial insecurity during the pandemic in Extreme Responders/Resisters and Sufferers. The finding of financial insecurity may be related to the worse coping scores also present in these two groups.

Furthermore, it can also be seen in Table 1 that between-clusters differences in our study are generally smaller than those described by Duffy and Allington [22], particularly with respect to our outcome variables of poor coping and adherence.

Excitingly, there is clear consistency in cluster attributes between these two studies, despite significant methodological differences. This external consistency lends strength to the assertion that three clear attribute sets can be distinguished in individual pandemic responses. Although the validity of the variance in adherence and poor coping explained by clusters is limited (as they were created with some elements in common), this is mitigated by the recurrence of three main groups in both analyses.

Based on the findings of this study, all demographic variables combined accounted for 15.9% of total variance in adherence and between-cluster differences accounted for an additional 10.7% of the variance. Therefore, policies aiming to improve adherence could target the Extreme Respondent cluster as well as demographic variables such as gender (men) and politics (those who identify as right-wing). Additionally, our data also supports the role of lockdown stringency on adherence. On the other hand, all demographic variables combined accounted for only 4.1% of total variance in coping scores, while between-cluster differences accounted for an additional 31.0% of the variance. This suggests that targeting clusters may be the most effective way to improve coping.

From this it can be seen that cluster analysis is valid technique to measure adversity responses, even in an international sample, and can reliably identify homogeneous groups within a broad dataset. That said, 73.5% of variability in adherence scores and 64.0% of variability in poor coping scores remained unexplained. This, combined with the finding that clustering appears to be more viable than demographics as a determinant of adversity responses, suggests that nuances in personality and environmental factors should be considered for identifying predictors of coping and adherence in future research. For example, individuals with high scores on the “Big Five” personality domain “Agreeableness” may be more inclined to comply with policy measures against COVID-19, while individuals with lower emotional stability tend to hoard supplies and fear financial [45, 46].

Other limitations of this study include low variability in responses suggests that the reliability and ecological validity of the self-reported data should be questioned. Furthermore, the heavy skew towards young individuals, and lack of representation of nonbinary genders (who may be at increased risk for adversity during pandemic restrictions), limit the generalisability of our findings across these dimensions. Data regarding self-identified race, culture and ethnicity was not collected, and collecting this information as well as clearer or more standardised measures of socio-economic status and level of education would enhance the quality of the results found here. To improve the validity of future cluster analyses for COVID-related decision making, the model would benefit by keeping variables used for cluster creation and variables used for outcome measures mutually exclusive.

In closing, each country has approached the COVID-19 pandemic in different ways, yet we demonstrate here that people have formed internationally coherent clusters in responding to this challenging period. Incorporating group-specific approaches to improve adherence and coping will not only assist in reducing the spread of infection, but also benefit the well-being of citizens while vaccinations are under way. This knowledge should inform way we create, enforce and adapt restrictive regulations for future large-scale crises.

## Data Availability

Data has been deposited at Dryad and will be made available on publication

https://doi.org/10.5061/dryad.qv9s4mwfc

## Supporting information captions

S1: A full list of the survey questions and answer options used for this study.

S2: A figure describing the inclusion and exclusion criteria and population size. Inclusion and exclusion criteria used. Statistical outliers were identified using Mahalanobis distance. Participants with invalid values for demographic variables were excluded due to the requirements of regression analyses. Numbers for excluded participants due to demographics may differ to those described in the Population Distribution section of the main text, as participants who met multiple exclusion criteria are reported in each demographic category in the main text, but are only included once in the figure above. SSF = Social security factor, NCRF = National coronavirus response factor.

S3: A diagrammatic overview of the variables and tests used in the study.

S4: A diagrammatic view of the variables included in each model for the hierarchical regression analyses in order to assess their relative effect on adherence and poor coping outcomes.

S5: The rotated component matrix for the principal component analysis of 54 lockdown questions, showing the factor loading for each lockdown question. An eigenvalue of 1, and varimax rotation with Kaiser normalisation were used. The rotation converged in 8 iterations, with 12 factors identified. Factor loadings above 0.5 (green) or below -0.5 (orange) are highlighted.

S6: A tabulated summary of the question themes of each lockdown factor, derived from a principal component analysis of responses for the 53 lockdown-related questions.

S7: The rotated component matrix from a principal component analysis of participants’ scores for 6 geopolitical indices based on participant country and date of response. Values show the factor loading for each index, using an eigenvalue of 1. Varimax Rotation with Kaiser Normalisation was used, and the rotation converged in 3 iterations, with 2 factors identified. Factor loadings > 0.5 are highlighted in green.

S8: Statistical outputs relevant for testing the assumptions of the hierarchical regressions to assess the determinants of adherence and poor coping.

## Declarations

### Ethics approval

The questionnaire and methodology for this study was approved by Liverpool John Moores University, UK Research Ethics Committee (UREC ref: 20/NSP/035).

### Consent to participate

Informed consent was obtained from all individual participants included in the study.

### Consent for publication

No identifying data or images of individual participants which would require additional consent for publication are included.

### Availability of data and code

Will be made available from Dryad at https://doi.org/10.5061/dryad.qv9s4mwfc on publication

### Authors’ contributions

S.J.B. conceptualised the project, was responsible for project administration and conducted the investigation. S.V.B curated the data, developed the methodology and software and performed formal analysis. A.D and S.V.B. prepared the original draft. S.J.B. and H.B.S. supervised S.V.B and A.D., enduring validity through verification and reviewing and editing throughout draft preparation. All authors approved the final version of the paper for submission.

---

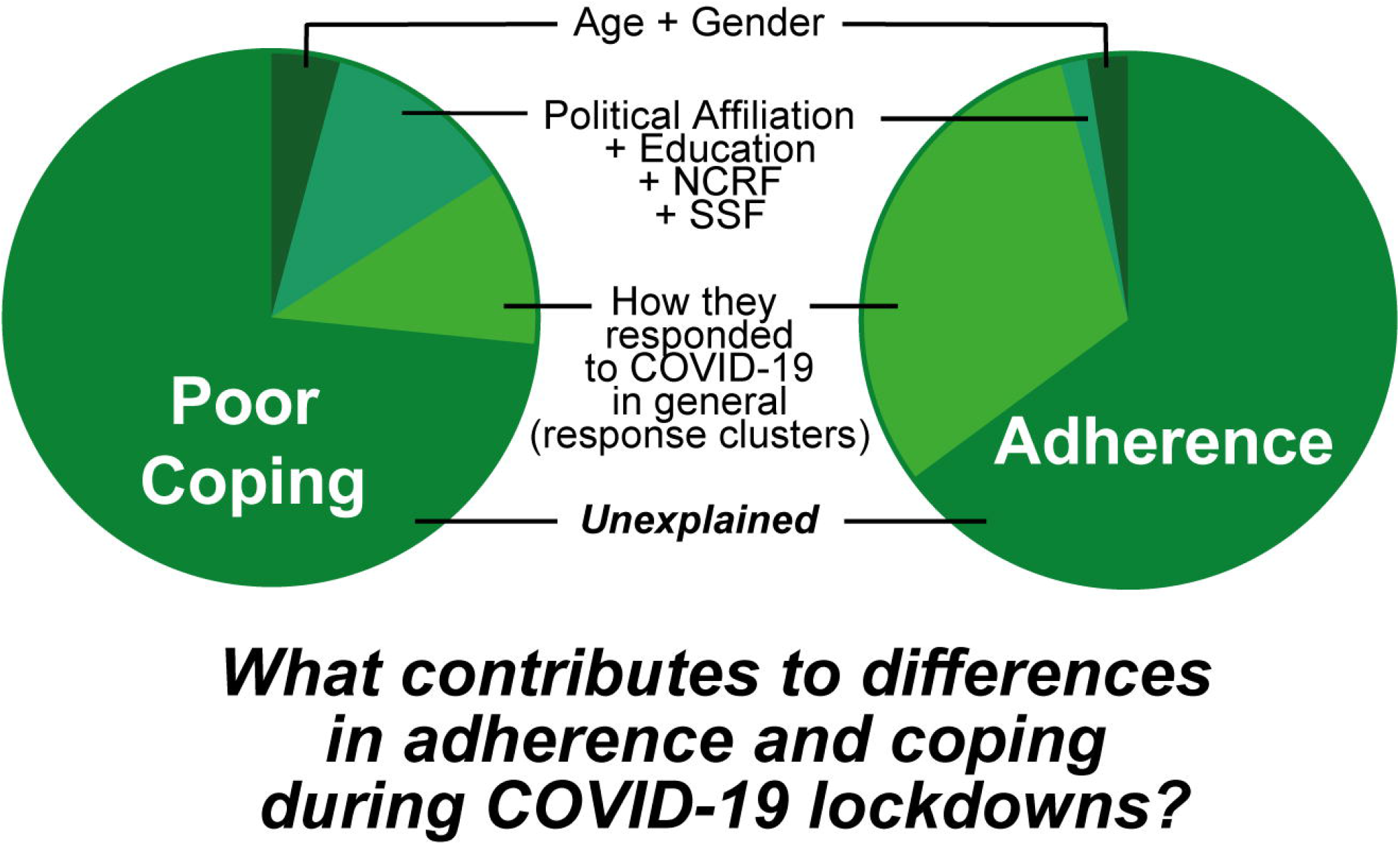

