## Supplementary material for "Profiling lockdown adherence and poor coping responses towards the COVID-19 crisis in an international cross-sectional survey": S1

### Survey Questions:

#### Demographic Questions:

What is your age? *Please note: do not continue if you are under 18 or over 75 years of age.

*Text field restricted to numerals*

What gender do you currently identify with?

*Multiple choice:*

- Male
- Female
- Prefer not to be identified by gender
- Other

Please write which country (including the region) which you are from in the box below:

*Free text*

On which side of the political divide do you tend to identify with?

*Multiple choice:*

- Far left
- Left
- Central
- Right
- Far right
- No political interest

What is your highest level of education? Please write in the box below (e.g. school-level exams, undergraduate degree, masters degree, PhD)

*Free text*

### Lockdown Questions:

Q1. I followed the COVID lockdown rules

- Completely
- Nearly all the time
- Some of the time
- Hardly ever
- None of the time

Q2. I supported lockdown measures

- Completely
- Nearly all the time
- Some of the time
- Hardly ever
- None of the time

Q3. I am adhering to social distancing rules

- Completely
- Nearly all the time
- Some of the time
- Hardly ever
- None of the time

Q4. I wash hands more often for at least 20 seconds

- Completely
- Nearly all the time
- Some of the time
- Hardly ever
- None of the time

Q5. I cover my mouth when coughing

- Completely
- Nearly all the time
- Some of the time
- Hardly ever
- None of the time

Q6. I avoid close contact with someone who is infected

- Completely
- Nearly all the time
- Some of the time
- Hardly ever
- None of the time

Q7. I have been avoiding places where many people are most likely to gather (e.g. parks, beaches, other outdoor spaces)

- Completely
- Nearly all the time
- Some of the time
- Hardly ever
- None of the time

Q8. I have been taking homeopathic remedies to help overcome the coronavirus

- Often
- Nearly all the time
- Some of the time
- Hardly ever
- None of the time

Q9. I have been taking herbal remedies to help overcome the coronavirus

- Often
- Nearly all the time
- Some of the time
- Hardly ever
- None of the time

Q10. I have been avoiding eating meat to help overcome the coronavirus

- Often
- Nearly all the time
- Some of the time
- Hardly ever
- None of the time

Q11. I have been drinking ginger tea to help overcome the coronavirus

- Often
- Nearly all the time
- Some of the time
- Hardly ever
- None of the time

Q12. I have been using antibiotics to help overcome the coronavirus

- Often
- Nearly all the time
- Some of the time
- Hardly ever
- None of the time

Q13. During the COVID lockdown I met up with friends or family outside the home

- Often
- Nearly all the time
- Some of the time
- Hardly ever
- None of the time

Q14. During the COVID lockdown had friends or family visit me at home

- Often
- Nearly all the time
- Some of the time
- Hardly ever
- None of the time

Q15. I have been outside when having coronavirus-like symptoms

- Often
- Nearly all the time
- Some of the time
- Hardly ever
- None of the time

Q16. I have had a confirmed case of coronavirus

- Yes
- No

Q17. I have NOT had a confirmed case of coronavirus, but I think I MIGHT have had it

- Yes
- No

Q18. I have been in contact with a counselling or support service

- Often
- Nearly all the time
- Some of the time
- Hardly ever
- None of the time

Q19. I support police powers during the COVID crisis

- Completely
- Nearly all the time
- Some of the time
- Hardly ever
- None of the time

Q20. I agree with the following statement: "Too much fuss is being made about the risk of coronavirus":

- Completely
- Nearly all the time

- Some of the time
- Hardly ever
- None of the time

Q21. I agree with the following statement: "The coronavirus was probably created in a laboratory":

- Completely
- Nearly all the time
- Some of the time
- Hardly ever
- None of the time

Q22. I agree with the following statement: "Most people in the UK have already had coronavirus without realising it":

- Completely
- Nearly all the time
- Some of the time
- Hardly ever
- None of the time

Q23. I agree with the following statement: "Pets can transmit coronavirus":

- Completely
- Nearly all the time
- Some of the time
- Hardly ever
- None of the time

Q24. I agree with the following statement: "Coronavirus can last on some surfaces for up to 7 days":

- Completely
- Nearly all the time
- Some of the time
- Hardly ever
- None of the time

Q25. I agree with the following statement: "Sanitising hand gels are more effective at protecting you from coronavirus than washing your hands with soap and water":

- Completely
- Nearly all the time
- Some of the time
- Hardly ever
- None of the time

Q26. I agree with the following statement: "The NHS recommends that you should wear a face mask when you are out, even if you do not have coronavirus":

- Completely
- Nearly all the time
- Some of the time
- Hardly ever
- None of the time

Q27. I agree with the following statement: "There will be a quick resolution to the coronavirus crisis and lockdown measures will end soon":

- Completely
- Nearly all the time
- Some of the time
- Hardly ever
- None of the time

Q28. I am closely following official guidance/recommendations on how to protect myself and others

- All recommendations
- Most recommendations
- Some recommendations

- Hardly any recommendations
- I am not following any of the recommendations

Q29. I have lost sleep over coronavirus
- Completely
- Worse than usual
- Some of the time
- Hardly ever
- None of the time

Q30. My eating patterns have changed during the coronavirus lockdown:
- Eating more than usual
- Eating less than usual
- Eating more or less the same as usual

Q31. I am eating much more HEALTHY food
- Definitely
- Somewhat
- Unsure
- Not really
- Definitely not

Q32. I am eating much more UNHEALTHY food
- Definitely
- Somewhat
- Unsure
- Not really
- Definitely not

Q33. I am drinking much more alcohol
- Definitely
- Somewhat
- Unsure
- Not really
- Definitely not

Q34. I am using non-prescription drugs (e.g. painkillers, other over-the-counter remedies) much more
- Definitely
- Somewhat
- Unsure
- Not really
- Definitely not

Q35. I am finding the coronavirus outbreak and/or the lockdown measures extremely difficult to cope with
- Definitely
- Somewhat
- Unsure
- Not really
- Definitely not

Q36. I have been spending time thinking about the coronavirus
- Completely
- Worse than usual
- Some of the time
- Hardly ever
- None of the time

Q37. I have argued more with family/people in the home during the COVID lockdown
- Completely
- Worse than usual
- Some of the time
- Hardly ever
- None of the time

Q38. I feel more anxious since the lockdown measures were introduced

- Completely
- Worse than usual
- Some of the time
- Hardly ever
- None of the time

Q39. I feel more depressed since the lockdown measures were introduced

- Completely
- Worse than usual
- Some of the time
- Hardly ever
- None of the time

Q40. I feel helpless as a result of coronavirus

- Completely
- Worse than usual
- Some of the time
- Hardly ever
- None of the time

Q41. I will lose my job as a result of coronavirus and/or the lockdown

- Certain
- Likely
- Unsure
- Unlikely
- Definitely not

Q42. I will experience financial difficulties as a result of coronavirus and/or the lockdown

- Certain
- Likely
- Unsure
- Unlikely
- Definitely not

Q43. Life will return to normal soon

- Certain
- Likely
- Unsure
- Unlikely
- Definitely not

Q44. The economy will start to grow again soon

- Certain
- Likely
- Unsure
- Unlikely
- Definitely not

Q45. We will be able to vaccinate the population against coronavirus soon

- Certain
- Likely
- Unsure
- Unlikely
- Definitely not

Q46. Schools will stay closed in the future

- Certain
- Likely
- Unsure
- Unlikely
- Definitely not

Q47. Older people and those with underlying health issues will continue to be asked to remain home

- Certain

- Likely
- Unsure
- Unlikely
- Definitely not

Q48. My country's government has handled the crisis:

- Consistently and organised
- Somewhat consistent and organised
- Unsure
- Somewhat inconsistent and confused
- Inconsistently and confused

Q49. I would describe my trust in my country's government to control the spread of coronavirus as:

- Certain
- Likely
- Unsure
- Unlikely
- Definitely not

Q50. I would describe my trust in the coronavirus information my country's government provides as:

- Certain
- Likely
- Unsure
- Unlikely
- Definitely not

Q51. My belief that the government acted too slowly to control the spread of coronavirus is:

- Certain
- Likely
- Unsure
- Unlikely
- Definitely not

Q52. My belief that the government communication provided helpful advice:

- Certain
- Likely
- Unsure
- Unlikely
- Definitely not

Q53. My belief that the government plan responded well to the changing scientific information and situation:

- Certain
- Likely
- Unsure
- Unlikely
- Definitely not

Q54. I check social media for information or updates about coronavirus:

- Daily or more frequently
- Once a day
- Once every few days
- Once a week
- Less than once a week or never
