## Supplementary figures and images for "Profiling lockdown adherence and poor coping responses towards the COVID-19 crisis in an international cross-sectional survey"

### S2

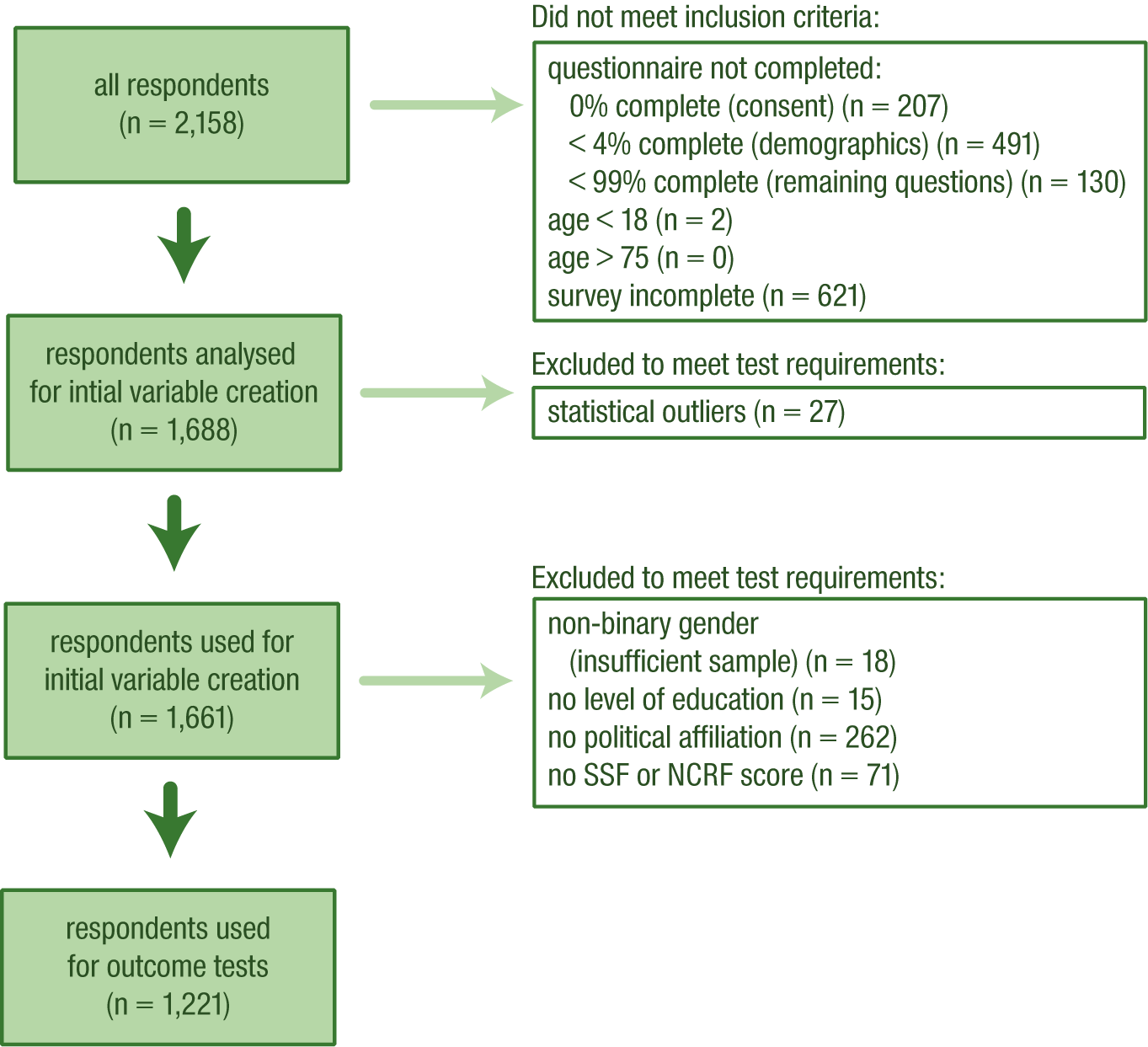

### S3

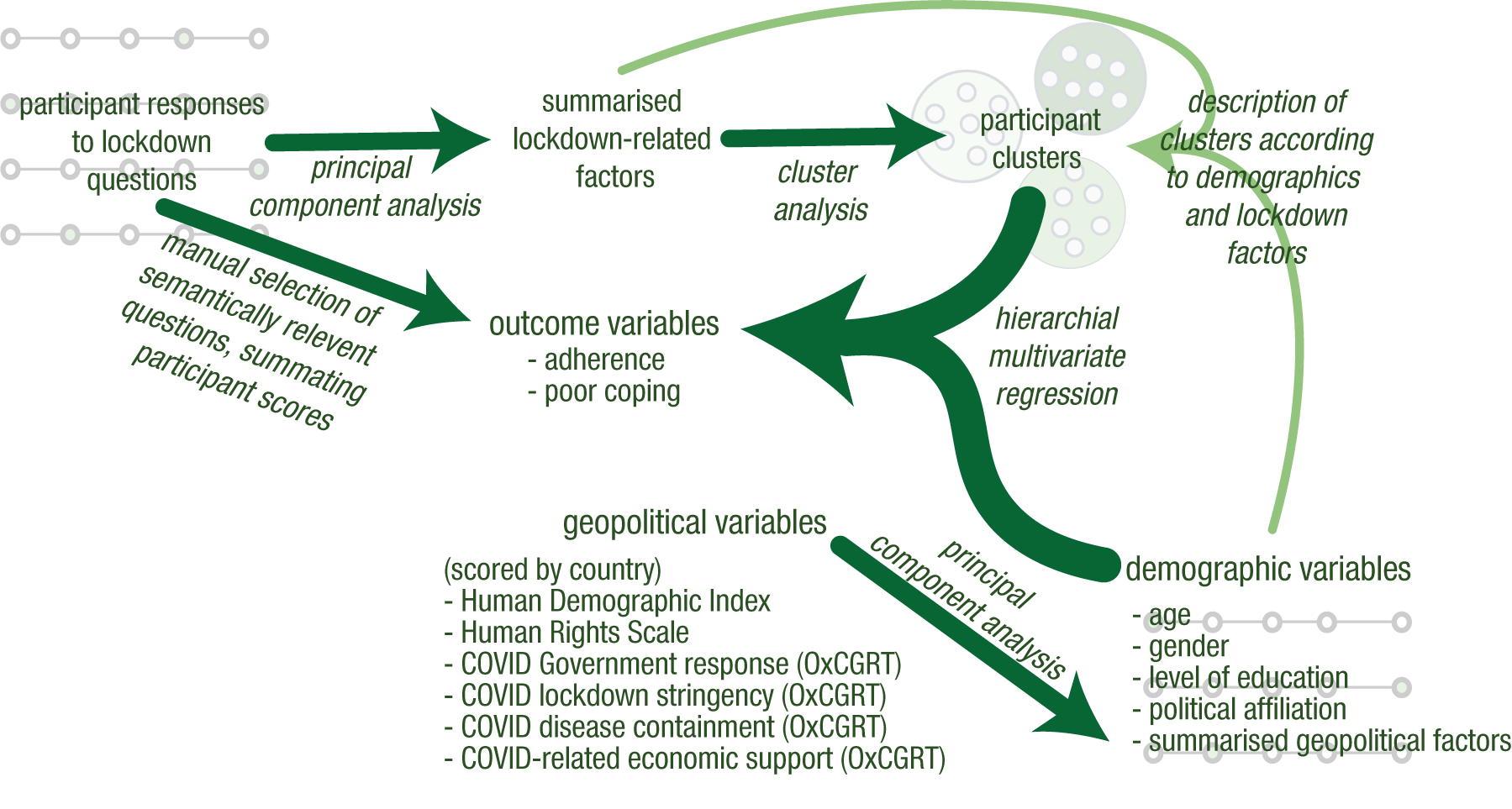

### S4

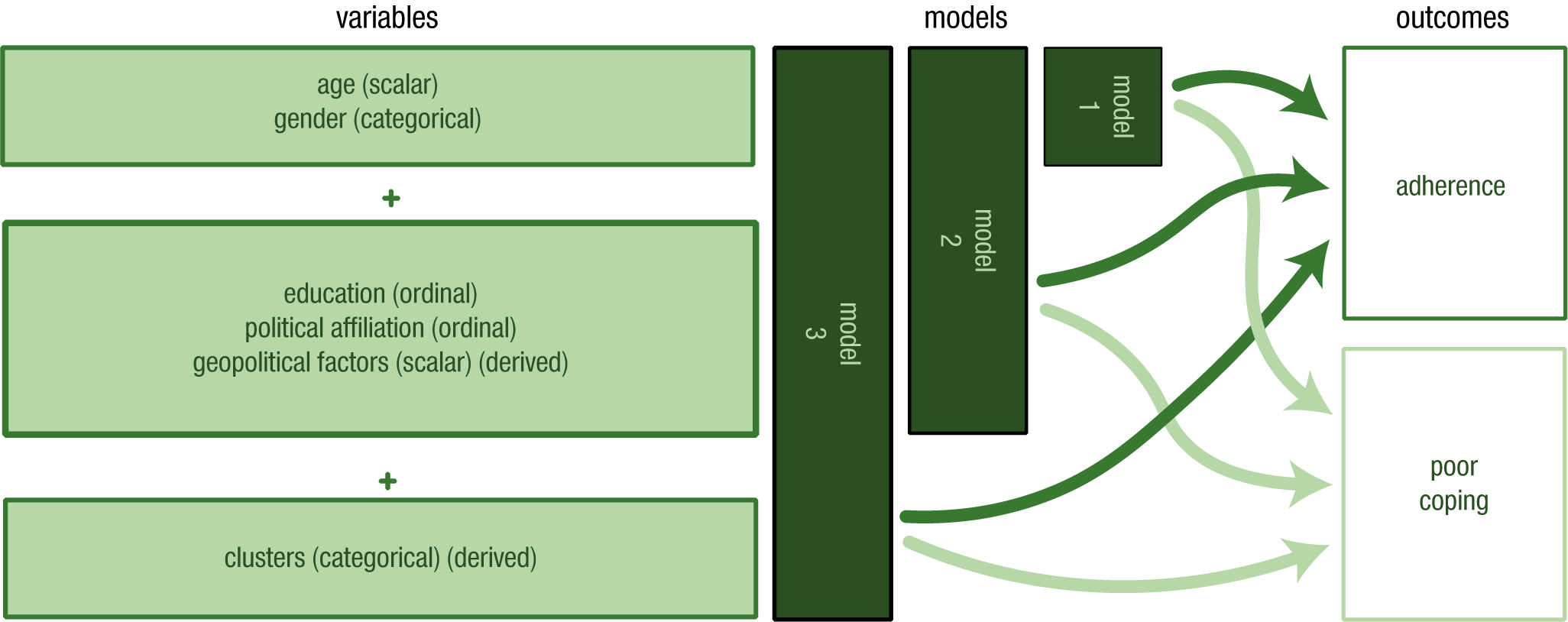
