## Supplementary material for "Profiling lockdown adherence and poor coping responses towards the COVID-19 crisis in an international cross-sectional survey": S5

| | Component | | | | | | | | | | | |
|---|---|---|---|---|---|---|---|---|---|---|---|---|
| | 1 | 2 | 3 | 4 | 5 | 6 | 7 | 8 | 9 | 10 | 11 | 12 |
| q1 | 0.098 | 0.11 | 0.059 | 0.631 | 0.004 | -0.07 | -0.07 | -0.18 | 0.042 | -0.02 | -0.05 | -0.11 |
| q2 | 0.042 | 0.173 | -0.01 | 0.627 | -0.02 | -0.27 | 0.106 | -0.17 | 0.07 | 0.139 | -0.09 | 0.042 |
| q3 | 0.1 | 0.055 | -0.04 | 0.719 | 0.016 | -0.07 | -0.04 | -0.08 | 0.007 | 0.025 | 0.05 | -0.04 |
| q4 | 0.075 | -0.07 | 0.089 | 0.559 | -0.06 | 0.173 | 0.091 | -0.02 | -0.04 | -0.08 | 0.025 | -0.08 |
| q5 | -0.24 | -0 | 0.074 | 0.55 | 0.031 | 0.178 | 0.03 | 0.1 | 0.059 | -0.17 | 0.027 | 0.138 |
| q6 | -0.28 | -0.02 | 0.019 | 0.539 | -0.03 | 0.085 | -0.09 | 0.23 | -0.12 | -0.05 | 0.005 | 0.093 |
| q7 | 0.128 | -0.03 | -0.03 | 0.637 | -0.01 | -0.08 | -0 | -0.09 | -0.04 | 0.07 | 0.019 | -0.18 |
| q8 | 0.767 | 0.036 | 0.073 | 0.036 | 0.113 | 0.182 | -0.01 | 0.047 | 0 | 0.036 | 0.022 | -0.06 |
| q9 | 0.776 | 0.019 | 0.089 | 0.035 | 0.1 | 0.196 | 0.004 | 0.029 | -0 | 0.021 | 0.053 | -0.07 |
| q10 | 0.676 | -0 | 0.07 | 0.046 | 0.091 | 0.064 | -0.05 | -0.04 | -0 | -0 | 0.083 | 0.096 |
| q11 | 0.765 | 0.048 | 0.107 | 0.012 | 0.092 | 0.13 | 0.008 | 0.096 | 0.048 | 0.043 | 0.049 | 0.037 |
| q12 | 0.749 | 0.079 | 0.089 | 0.023 | 0.076 | 0.096 | 0.093 | 0.216 | 0.051 | 0.086 | 0.008 | 0.042 |
| q13 | 0.209 | 0.009 | 0.016 | -0.24 | 0.173 | 0.08 | 0.076 | 0.741 | 0.042 | 0.032 | -0.01 | -0.01 |
| q14 | 0.309 | -0.02 | -0.01 | -0.21 | 0.114 | 0.067 | 0.084 | 0.723 | 0.127 | 0.01 | -0.01 | -0.03 |
| q15 | 0.598 | 0.115 | 0.125 | -0.18 | 0.087 | 0.004 | 0.091 | 0.319 | 0.015 | 0.12 | 0.024 | 0.198 |
| q16 | 0.29 | 0.039 | 0.081 | 0.004 | -0.01 | 0.049 | 0.005 | 0.148 | 0.127 | 0 | -0.04 | 0.391 |
| q17 | 0.096 | -0.07 | 0.091 | -0.1 | 0.041 | 0.148 | 0.091 | -0.11 | 0.001 | -0.02 | 0.097 | 0.682 |
| q18 | 0.498 | 0.007 | 0.21 | -0.01 | -0.01 | -0.07 | 0.003 | 0.111 | 0.131 | -0.04 | -0.01 | 0.334 |
| q19 | -0.02 | 0.214 | -0.03 | 0.321 | -0.02 | -0.15 | 0.096 | 0.004 | -0.28 | 0.299 | -0.1 | 0.306 |
| q20 | 0.178 | -0.04 | 0.021 | -0.31 | 0.192 | 0.546 | -0.04 | 0.218 | -0.07 | -0.08 | 0.074 | 0.059 |
| q21 | 0.226 | 0.008 | 0.043 | -0.06 | -0.01 | 0.66 | 0.023 | 0.028 | -0.01 | 0.127 | -0.01 | 0.016 |
| q22 | 0.17 | 0.002 | 0.065 | -0.03 | 0.063 | 0.672 | 0.091 | -0.05 | 0.001 | 0.191 | -0.01 | 0.184 |
| q23 | 0.173 | 0 | 0.021 | -0.03 | 0.017 | 0.125 | -0.04 | 0.133 | -0.04 | 0.715 | 0.077 | -0.1 |
| q24 | 0.047 | -0.03 | 0.044 | 0.106 | 0.068 | 0.155 | 0.011 | -0.09 | 0.108 | 0.691 | -0.01 | 0.066 |
| q25 | 0.184 | 0.134 | -0.01 | -0.06 | 0.189 | 0.334 | 0.142 | 0.126 | 0.223 | 0.214 | -0.08 | -0.17 |
| q26 | -0.14 | 0.082 | 0.014 | 0.477 | 0.012 | -0.28 | 0.057 | -0.11 | 0.241 | 0.199 | -0 | 0.197 |
| q27 | 0.309 | 0.156 | 0.06 | -0 | 0.576 | 0.293 | 0.029 | 0.052 | 0.012 | 0.066 | 0.05 | -0.06 |
| q28 | -0.08 | 0.112 | 0.038 | 0.617 | 0.04 | -0.25 | -0.07 | -0.13 | 0.109 | 0.157 | 0.012 | 0.125 |
| q29 | 0.225 | -0.03 | 0.642 | 0.1 | 0.017 | -0.09 | 0.139 | -0.04 | -0.05 | 0.094 | 0.01 | 0.033 |
| q30 | -0.11 | -0.01 | 0.022 | 0.102 | 0.001 | 0.046 | 0.668 | 0.013 | 0.093 | -0.02 | -0.04 | 0.12 |
| q31 | 0.391 | 0.041 | 0.145 | 0.131 | 0.164 | 0.05 | -0.34 | -0.14 | 0.076 | 0.083 | 0.042 | 0.069 |
| q32 | 0.08 | -0.05 | 0.264 | -0.01 | 0.047 | 0.017 | 0.728 | 0.013 | 0.057 | 0.044 | 0.101 | -0 |
| q33 | 0.259 | 0.011 | 0.158 | -0.09 | 0.017 | 0.057 | 0.563 | 0.091 | -0.15 | -0.01 | 0.099 | -0 |
| q34 | 0.526 | 0.015 | 0.296 | -0.11 | 0.067 | 0.031 | 0.292 | -0.02 | -0.04 | 0.119 | 0.048 | -0.01 |
| q35 | 0.104 | -0.08 | 0.673 | -0.11 | -0.04 | 0.07 | 0.078 | 0.094 | -0.09 | 0.019 | 0.059 | 0.032 |
| q36 | 0.109 | 0.048 | 0.636 | 0.222 | -0.03 | -0.04 | -0.03 | 0.001 | 0.021 | 0.06 | -0.01 | 0.164 |
| q37 | 0.113 | 0.074 | 0.534 | -0.04 | 0.123 | 0.106 | 0.11 | -0.04 | 0.241 | -0.09 | 0.085 | 0.031 |
| q38 | 0.05 | -0.03 | 0.825 | 0.057 | -0.01 | 0.052 | 0.008 | -0 | 0.001 | -0.02 | 0.065 | 0.004 |
| q39 | 0.013 | -0.03 | 0.796 | -0.02 | -0.04 | 0.05 | 0.064 | 0.037 | 0.027 | -0.03 | 0.158 | 0.015 |
| q40 | 0.138 | -0.07 | 0.763 | -0.03 | -0.06 | -0.01 | 0.045 | -0.02 | 0.018 | 0.037 | 0.169 | -0.02 |
| q41 | 0.228 | -0.02 | 0.238 | -0.03 | -0.04 | 0.001 | 0.094 | -0.01 | 0.008 | 0.033 | 0.78 | 0.025 |
| q42 | 0.029 | -0.05 | 0.326 | 0.059 | -0.07 | 0.02 | 0.05 | 0.003 | 0.141 | 0.037 | 0.773 | 0.039 |
| q43 | 0.136 | 0.15 | -0.07 | -0.07 | 0.805 | 0.137 | -0.02 | 0.097 | -0.05 | -0.06 | -0.06 | -0.02 |
| q44 | 0.09 | 0.155 | -0.06 | -0.01 | 0.791 | 0.023 | 0.002 | 0.095 | -0.1 | 0.087 | -0.04 | 0.046 |
| q45 | 0.108 | 0.144 | 0.028 | 0.028 | 0.739 | -0.1 | 0.025 | 0.02 | 0.077 | 0.023 | -0.03 | 0.036 |
| q46 | 0.212 | -0.03 | 0.06 | -0.04 | -0.04 | -0.03 | 0.062 | 0.068 | 0.715 | -0 | 0.006 | -0.03 |
| q47 | -0.13 | 0.031 | -0.03 | 0.163 | -0.06 | -0.04 | -0.08 | 0.08 | 0.641 | 0.067 | 0.15 | 0.151 |
| q48 | 0.054 | 0.868 | 0.012 | 0.052 | 0.125 | 0.056 | -0.01 | 0.026 | 0.036 | -0.01 | -0.06 | -0.05 |
| q49 | 0.116 | 0.881 | -0.02 | 0.034 | 0.137 | 0.02 | 0.034 | -0.02 | -0.02 | 0.029 | -0.03 | -0.02 |
| q50 | 0.082 | 0.83 | 0.039 | 0.075 | 0.075 | -0.11 | 0.046 | 0.005 | -0.02 | 0.032 | -0.03 | 0.052 |
| q51 | 0.086 | -0.52 | 0.127 | 0.162 | -0.02 | -0.13 | 0.104 | -0.07 | -0.08 | 0.242 | 0.046 | 0.188 |
| q52 | 0.012 | 0.797 | -0.05 | 0.106 | 0.119 | -0.07 | -0.03 | -0.03 | -0.01 | 0.027 | 0.061 | 0.077 |
| q53 | 0.038 | 0.828 | -0.03 | 0.11 | 0.106 | 0.043 | -0.04 | -0.03 | -0 | 0.035 | 0.004 | 0.026 |
