## Supplementary material for "Profiling lockdown adherence and poor coping responses towards the COVID-19 crisis in an international cross-sectional survey": S6

| Factor alias | Questions included in factor variables (factor loading >0.5 or < -0.5): |
|---|---|
| self-treatment | Questions 8-12 and 34.<br>A score near 5 suggests that the participant had engaged on average in most of the following behaviours to "overcome the coronavirus" **often**:<br>*taken homeopathic remedies, taken herbal remedies, avoided meat, drunk ginger tea, used antibiotics and used more non-prescription drugs* |
| government response | Questions 48-50, 52, 53<br>A score near 5 suggests that the participant was on average **certain** about most of the following:<br>*they believed their government's handling of the crisis was consistent and organised, they trusted their government to control the spread of coronavirus, they trusted the information provided by the government, they believed their government had not acted too slowly to control the spread of COVID, that the communication from their government was helpful and that their government responded well to the changing scientific information and situation.* |
| poor mood | Questions 36-40<br>A score near 5 suggests that the participant felt that most of the following was on average **completely** true since lockdown measures were introduced:<br>*they have been spending time thinking about the coronavirus, they have argued more at home, they have been more anxious, they have felt more depressed and they have felt helpless* |
| adherence | Questions 1-7, 28<br>A score near 5 suggests that the participant felt that most of the following was on average **completely** true:<br>*they have followed lockdown rules, supported lockdown measures, adhered to social distancing rules, washed their hands more often for at least 20 seconds, covered their mouth when coughing, avoided close contact with infected individuals, avoided places where people are likely to gather and closely followed official guidance/recommendations on how to protect themselves and others.* |
| fringe beliefs | Questions 20-22<br>A score near 5 suggests that the participant agreed on average **completely** with most of the following:<br>*that too much fuss was being made about the risk of coronavirus, that it was probably created in a laboratory, and that most people in the UK had probably already had coronavirus without realising it.* |
| optimism | Questions 27, 43-45<br>A score near 5 suggests that the participant agreed on average **completely** with most of the following:<br>*there would be a quick resolution to the coronavirus crisis resulting in the end of lockdown measures, life would return to normal soon, the economy would soon begin to grow again and they would soon be able to vaccinate the population*. |
| unhealthy consumption | Questions 30, 32, 33<br>A score near 5 suggests that the participant agreed on average **completely** with most of the following:<br>*they were eating more than usual, they were eating more unhealthy food, and they were drinking more alcohol*. |
| in-person contact | Questions 13, 14<br>A score near 5 suggests that the participant engaged in these behaviours on average relatively **often** during the COVID lockdown:<br>*met up with friends or family outside the home, and had friends or family visit them.* |
| lockdown expectations | Questions 46, 47<br>A score near 5 suggests that the participant was on average **certain** that:<br>*schools would stay closed in the future and that older people and those with underlying health issues would continue to be asked to remain home* |
| indirect transmission | Questions 23, 24<br>A score near 5 suggests that the participant agreed on average **completely** with most of the following:<br>*pets can transmit coronavirus and coronavirus can last on some surfaces for up to 7 days":* |
| financial insecurity | Questions 41, 42<br>A score near 5 suggests that the participant was on average **certain** that, as a result of the coronavirus and/or the lockdown:<br>*they would lose their job and experience financial difficulties.* |
| suspected infection | Question 17<br>A score of 2 indicates that the participant **believed** on average that *they had not had a confirmed case of coronavirus, but that they might have had it*, while a score of 1 indicates the inverse. |

Questions not included in factors:
Q15. I have been outside when having coronavirus-like symptoms
Q16. I have had a confirmed case of coronavirus
Q18. I have been in contact with a counselling or support service
Q19. I support police powers during the COVID crisis
Q25. I agree with the following statement: "Sanitising hand gels are more effective at and water": coronavirus than washing your hands with soap protecting you from
Q26. I agree with the following statement: "The NHS recommends that you should wear a face mask when you are out, even if you do not have coronavirus"
31. I am eating much more HEALTHY food
54. I check social media for information or updates about coronavirus
