## Supplementary material for "Profiling lockdown adherence and poor coping responses towards the COVID-19 crisis in an international cross-sectional survey": S7

| | Component | |
|---|---|---|
| | 1 | 2 |
| HRS | -0.189 | 0.876 |
| HDI | -0.022 | 0.879 |
| OxCGRT: government response | 0.983 | 0.168 |
| OxCGRT: economic support | 0.547 | 0.734 |
| OxCGRT: containment and health policies | 0.986 | -0.088 |
| OxCGRT: lockdown stringency | 0.967 | -0.122 |
