## Supplementary material for "Profiling lockdown adherence and poor coping responses towards the COVID-19 crisis in an international cross-sectional survey": S8

### Regression assumptions:

- Adherence:
  - linear on assessment of partial regression plots and a plot of studentized residuals against the predicted values.
  - Independence of residuals: Durbin-Watson statistic of 1.94.
  - Homoscedastic: visual inspection of a plot of studentized residuals versus unstandardised predicted values.
  - No evidence of multicollinearity: largest VIF at 1.51.
  - Outliers: 13 cases with studentized deleted residuals < ±3 standard deviations, however, they were not excluded as their influence was likely low:
    - Leverage values: all below the “safe” value of 0.2 (highest = 0.021)
    - Cook’s values: all well below 1 (highest = 0.050)
    - No cases were not excluded from the regression as their influence was likely low.
  - The assumption of normality was met using a frequency histogram and a P-P plot of standardised residuals.
- Poor coping
  - linear on assessment of partial regression plots and a plot of studentized residuals against the predicted values.
  - Independence of residuals: Durbin-Watson statistic of 2.024.
  - Homoscedastic: visual inspection of a plot of studentized residuals versus unstandardised predicted values.
  - No evidence of multicollinearity: largest VIF at 1.51.
  - Outliers: 7 cases with studentized deleted residuals < ±3 standard deviations, however, they were not excluded as their influence was likely low:
    - Leverage values: all below the “safe” value of 0.2 (highest = 0.021)
    - Cook’s values: all well below 1 (highest = 0.019)
    - No cases were not excluded from the regression as their influence was likely low.
  - The assumption of normality was met using a frequency histogram and a P-P plot of standardised residuals.
  - Poor coping and adherence did not correlate on an independent regression analysis, with $p = .063$

*Table: Table show correlation coefficients (green saturation corresponds to strength of association) and corresponding p-values (red = not significant) for correlations between each independent variable. Extr = extreme responers. Suff = sufferers*

| Correlations between independent variables used in regressions | | | | | | | | | |
|---|---|---|---|---|---|---|---|---|---|
| | | age | gender | politics | education | NCRF | SSF | extr | suff |
| Pearson coefficient | age | 1.000 | -0.112 | 0.063 | 0.186 | 0.187 | 0.254 | -0.071 | 0.030 |
| | Gender | -0.112 | 1.000 | 0.236 | -0.077 | -0.156 | -0.266 | 0.182 | -0.186 |
| | politics | 0.063 | 0.236 | 1.000 | -0.068 | -0.137 | -0.161 | 0.220 | -0.197 |
| | Education | 0.186 | -0.077 | -0.068 | 1.000 | 0.185 | 0.063 | 0.005 | 0.009 |
| | NCRF | 0.187 | -0.156 | -0.137 | 0.185 | 1.000 | 0.520 | -0.118 | 0.107 |
| | SSF | 0.254 | -0.266 | -0.161 | 0.063 | 0.520 | 1.000 | -0.235 | 0.171 |
| | extr | -0.071 | 0.182 | 0.220 | 0.005 | -0.118 | -0.235 | 1.000 | -0.304 |
| | suff | 0.030 | -0.186 | -0.197 | 0.009 | 0.107 | 0.171 | -0.304 | 1.000 |
| Significance | age | . | 0.000 | 0.014 | 0.000 | 0.000 | 0.000 | 0.006 | 0.147 |
| | Gender | 0.000 | . | 0.000 | 0.003 | 0.000 | 0.000 | 0.000 | 0.000 |
| | politics | 0.014 | 0.000 | . | 0.009 | 0.000 | 0.000 | 0.000 | 0.000 |
| | Education | 0.000 | 0.003 | 0.009 | . | 0.000 | 0.013 | 0.433 | 0.378 |
| | NCRF | 0.000 | 0.000 | 0.000 | 0.000 | . | 0.000 | 0.000 | 0.000 |
| | SSF | 0.000 | 0.000 | 0.000 | 0.013 | 0.000 | . | 0.000 | 0.000 |
| | extr | 0.006 | 0.000 | 0.000 | 0.433 | 0.000 | 0.000 | . | 0.000 |
| | suff | 0.147 | 0.000 | 0.000 | 0.378 | 0.000 | 0.000 | 0.000 | . |

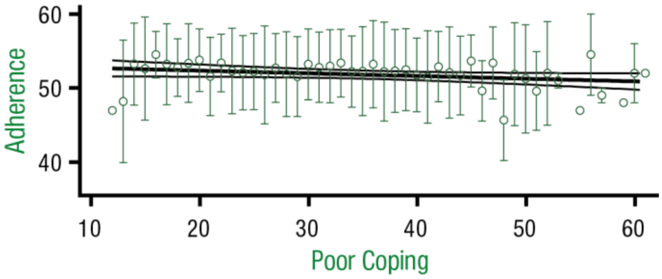


*Figure: Regression line when adherence score for each integer score for Poor Coping (error bars show mean and standard deviation), showing that there is no correlation is shown between adherence and poor coping. The regression equation is* $y = -0.0361x + 53.1$*, with an* $R^2 = 0.071$ *and P = 0.063.*

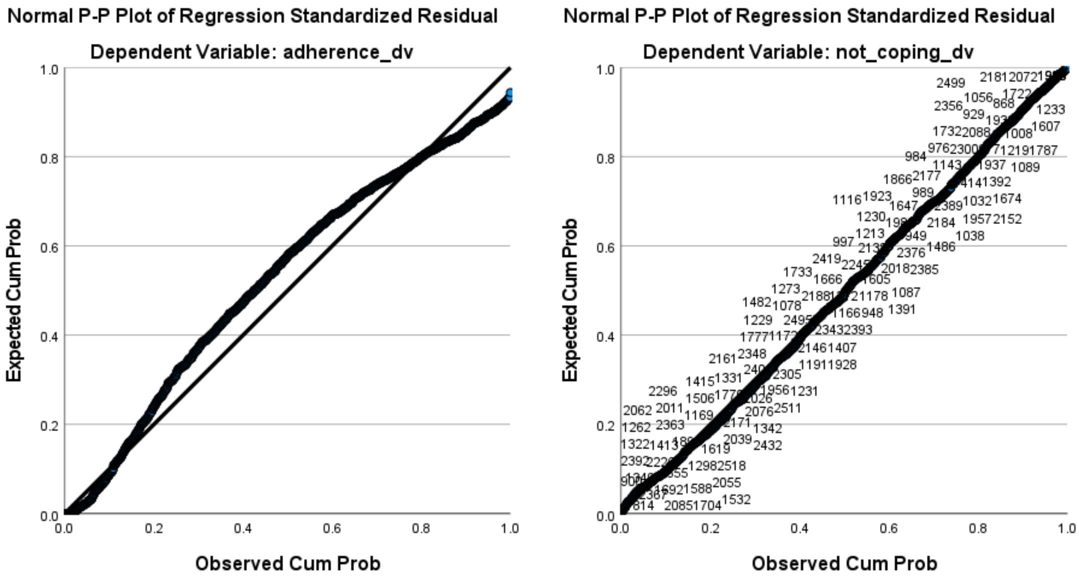


*Figure: P-P plots for assessing distribution of variance in adherence regression scores generated in SPSS from analysis output. The plot for adherence is on the left, and the plot for poor coping is on the right. Cum. = cumulative. Prob = Probability*
